# Identifying molecular pathways of type 2 diabetes using proteomics, metabolic, and anthropometric profiles in UK and Chinese adults

**DOI:** 10.64898/2025.12.19.25342701

**Authors:** Junxi Liu, Lingyan Chen, Reka Nagy, Neil Roberston, Matthew Traylor, Alfred Pozarickij, Lazaros Belbasis, Saredo Said, Wei Gan, Goutham Alta, Iona Millwood, Robin Walters, Huaidong Du, Pang Yao, Jun Lv, Canqing Yu, Dianjianyi Sun, Pei Pei, Liming Li, Zhengming Chen, Joanna M M Howson

## Abstract

**Introduction:** Proteogenomic analyses in large biobanks provide opportunities to improve understanding of the aetiology of type 2 diabetes (T2D) and to identify potential therapeutic targets.

**Methods:** We identified proteins (Olink Explore) associated with glycaemic traits and/or T2D with observational designs in UK Biobank (UKB-EUR, n =33,301). Bayesian non-negative matrix factorisation (bNMF) was applied to cluster T2D-associated proteins incorporating their phenotypic associations with 43 metabolic/anthropometric traits. For clusters’ leading proteins (top 10% by ranking), two-steps colocalization and bidirectional Mendelian randomization were performed to investigate three-way (i.e., protein–metabolic/anthropometric traits–T2D) relationships. Equivalent genetic analyses were conducted in the China Kadoorie Biobank (CKB-EAS, n = 2,029) to evaluate shared and ancestry-specific findings.

**Results:** A total of 1,793 proteins were observationally associated with glycaemic traits and/or T2D in UKB-EUR. Using bNMF, these proteins were classified into five clusters (Adiposity, Reduced adiposity, Lipids, Liver, and Kidney), of which the Reduced adiposity and Kidney clusters were novel; 906 proteins were identified as cluster-leading. Triangulation of observational and genetic evidence identified five proteins (B4GAT1, DNER, ENO3, HMOX2, OMG) potentially affecting T2D in UKB-EUR, one (ENTR1) in CKB-EAS, and three (RTBDN, TSPAN8, NCR3LG1) in both populations. In UKB-EUR, six proteins (CD34, FGFBP3, GALNT10, KHK, MENT, MXRA8) appeared to be consequences of T2D, while five proteins (GSTA1, GSTA3, MEGF9, NCAN, SHBG) showed bidirectional associations with T2D. Genetic analyses also suggested potential mechanistic pathways in T2D aetiology, including effects of RTBDN and TSPAN8 on T2D mediated through BMI and SHBG, respectively.

**Conclusions:** We identified multiple candidate proteins and biological pathways involved in T2D development, including novel protein clusters reflecting disease heterogeneity. These findings improve understanding of the molecular architecture of T2D and highlight potential biomarkers and therapeutic targets that may support precision prevention and treatment strategies.

**Research insights:** *What is currently known about this topic?:*

- Current T2D therapies do not address disease heterogenous yielding different responses between patients.
- Plasma proteins offer opportunities to characterise T2D molecular biology.

*What is the key research question?:*

- Can T2D-associated proteins be clustered by phenotypic protein - risk factor associations?
- Are proteins within these phenotype-derived clusters causally associated with T2D?
- How do proteins link T2D through cluster-specific markers revealing the heterogeneous aetiology?

*What is new?:*

- Five protein clusters identified (Adiposity, Reduced-Adiposity, Lipids, Liver, Kidney); Reduced-Adiposity and Kidney clusters are novel.
- 20 causal proteins identified (19 EUR, 4 EAS), with shared cross-ancestry signals.

*How might this study influence clinical practice?:*

- Identifies protein drug targets for T2D and supports precision treatment via cluster mechanisms.

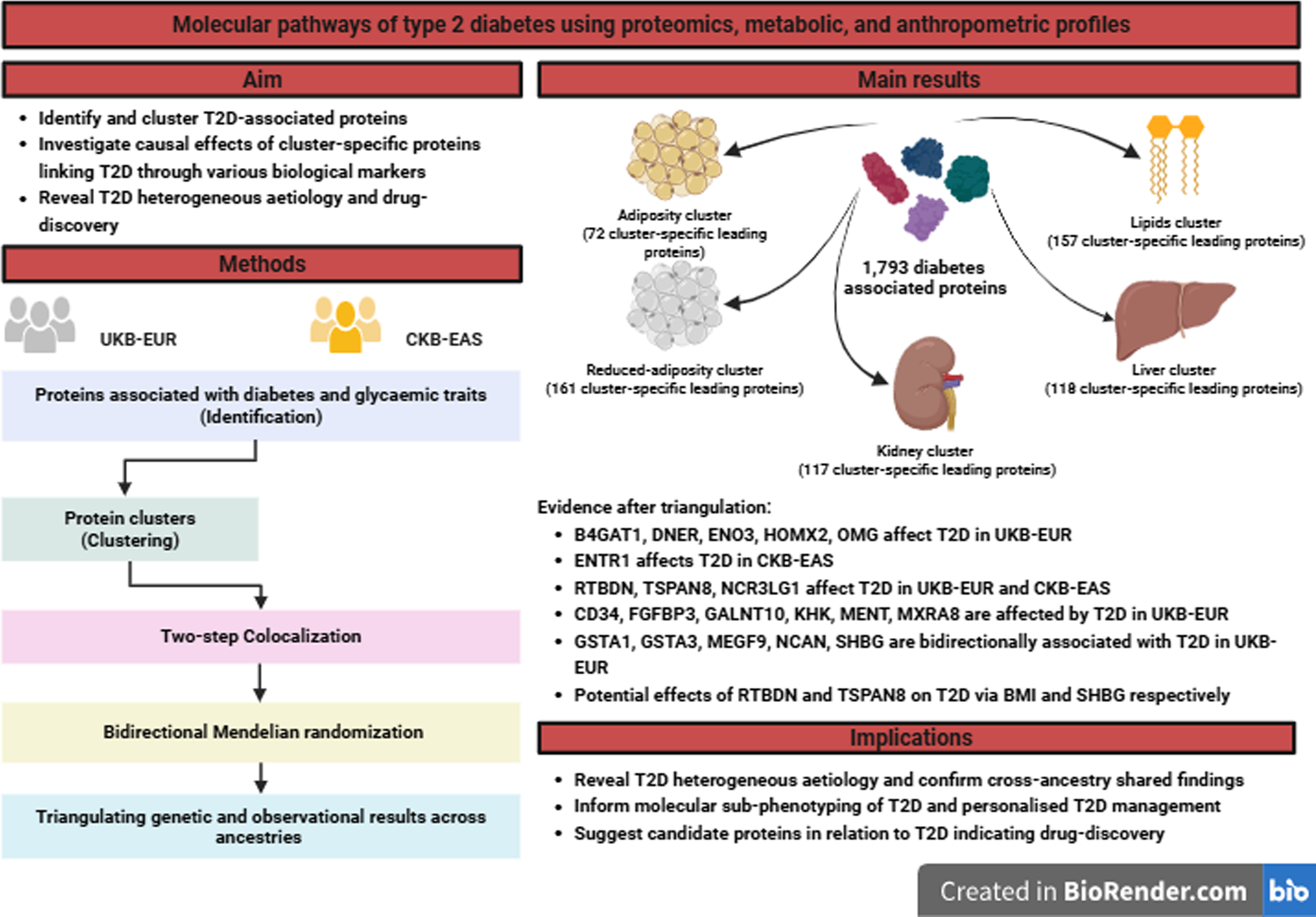

## Introduction

Approximately 10% of people worldwide will develop type 2 diabetes (T2D) by 2050, affecting about 1.3 billion people,[1] which constitutes a substantial health and socioeconomic burden.[2] Currently, T2D is treated and managed as a single disease though “individualized” treatments have been suggested based on different patient profiles (e.g., glycaemic, weight, risk for hypoglycemia, and history of cardiovascular, kidney, liver, and other comorbidities) and settings (e.g., children, older adults, pregnancy status).[3] To better characterise the aetiology of T2D and inform more targeted interventions and treatments, various strategies have been applied to identify T2D subtypes/clusters based on clinical features[4, 5] and/or genetic profile.[6–9] These have revealed heterogeneous T2D aetiology with potentially differing molecular subtypes (e.g., obesity, insulin, and dyslipidemia-related subtypes). Despite the promise, few studies have used additional omics data to further elucidate the molecular causes of T2D to better understand aetiology, improve risk prediction, and develop more personalised treatment of T2D.

In recent years, large-scale proteomics have been applied in population and clinical studies in diverse populations, which measures many thousands of proteins (including enzymes, antibodies, transport, and structural proteins) circulating in blood through active secretion, cellular leakages and damaged membrane.[10, 11] In addition to clinical and genetic information, proteomic data may provide new insights into T2D aetiology and molecular subtypes. Additionally, proteins are the targets of most drugs (e.g., glucagon-like peptide-1 receptor agonists[12] and sodium-glucose transport protein 2 inhibitors[13]) and may make attractive biomarkers for use in clinical development. Integrating genetics with proteomics can help identify novel T2D pathways and investigate causal links between proteins and T2D, relevant to drug discovery.[14–16]

In this cross-ancestry study of European (EUR) and East Asian population (EAS) study, we identified proteins associated with diabetes in UK Biobank (UKB-EUR). We classified/clustered these proteins based on their phenotypic associations with additional metabolic/anthropometric profiles in UKB-EUR. With genetic approaches, we identified three-way causal relationships (i.e., protein – metabolic/anthropometric traits – diabetes traits) in both UKB-EUR and China Kadoorie Biobank (CKB-EAS) to explore the shared and distinct findings between the two populations. Finally, we triangulated observational and genetic evidence across ancestries to identify the candidate proteins causally associated with T2D (**Figure 1**).

**Fig. 1.**
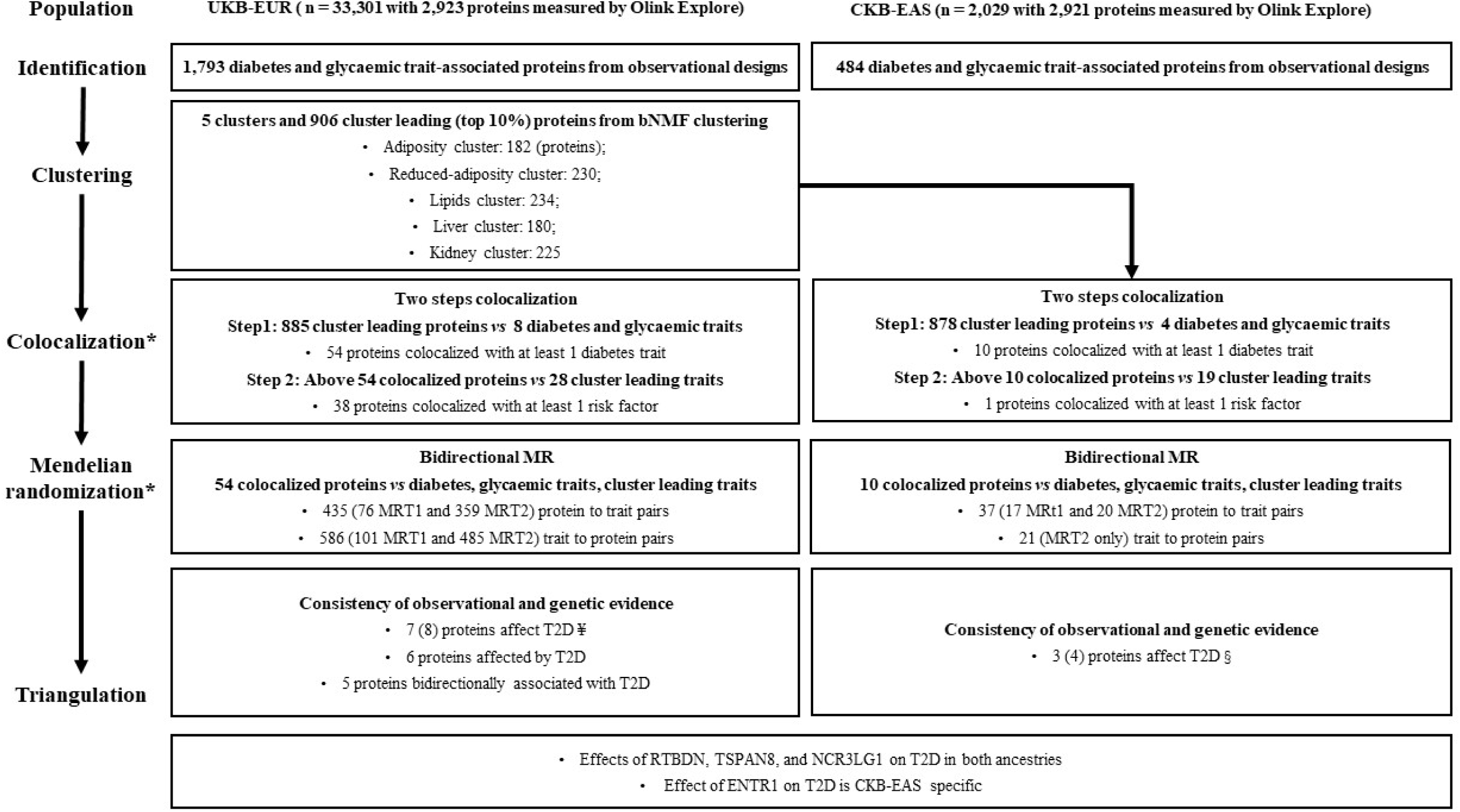
The flowchart of the study design, analytic approaches, and key finding. There was no metabolic trait information of the participants with proteins measured in CKB-EAS. The clustering step was not conducted. The colocalization analyses were conducted in the CKB-EAS with the proteins identified in the clustering step from UKB-EUR. * For colocalization, genetic variants in a +/-500kb window of the protein-coding gene were applied; in UKB-EUR, 21 proteins were encoded on the X chromosome, which were not processed to the genetic analyses in below due to no summary statistics of the X chromosome were available in the GWAS; in CKB-EAS, 28 proteins were coded on the X chromosome (21) or did not have pQTL in the CKB-EAS study and were excluded. For Mendelian randomization, the independent (R2 < 0.01 (UKB-EUR) / 0.05 (CKB-EAS), and p-value < 5E-08) cis-pQTLs in the protein regions were applied (Method). ¥ Apart from the 7 proteins (B4GAT1, DNER, ENO3, HMOX2, OMG, RTBDN, and TSPAN8) have consistent statistically robust observational and MR estimates, for protein NCR3LG1, the observational and genetic estimates were directionally consistent in both UKB-EUR and CKB-EAS though the observational estimate with iT2D in UKB-EUR crossed the null. We still considered it with robust evidence of an effect on T2D. § Apart from the 3 proteins (RTBND, NCR3LG1, and ENTR1) of the observational and Mendelian randomization estimates with T2D were statistically robust in CKB-EAS, the observational and genetic estimates of protein TSPAN8 with T2D were directionally consistent in both UKB-EUR and CKB-EAS, though the observational estimate with iT2D in CKB-EAS crossed the null. We still considered it with robust evidence of an effect on T2D. CKB-EAS: China Kadoorie Biobank East Asian population, UKB-EUR: UK Biobank European population, T2D: type 2 diabetes, bNMF: Bayesian non-negative matrix factorisation, MR: Mendelian randomization, TSPAN8: Tetraspanin-8, RTBDN: Retbindin, NCR3LG1: Natural cytotoxicity triggering receptor 3 ligand 1, ENTR1: Endosome-associated-trafficking regulator 1

## Methods

### Study population and Data

#### UK Biobank

The UK Biobank recruited 503,317 participants out of 9.2 million (5·5% response rate) eligible adults, aged between 40 and 69 years, in the UK from 2006 to 2010.[17, 18] At recruitment, a wide range of data were collected by questionnaire and physical measurements for health research after obtaining the participants’ informed consent.

Information on socio-demographic (e.g., education, deprivation index) and lifestyle (e.g., smoking, alcohol drinking, diet, physical activity) characteristics were obtained using a touchscreen questionnaire at the baseline assessment. For physical activity the summed metabolic equivalent task (MET) score per week for all activity was derived according to International Physical Activity Questionnaire (IPAQ) guidelines. For education, participants were asked which qualifications they had, which were transformed to five International Standard Classification of Education (ISCED) codes based on the years of education.[19] Townsend deprivation index[20] was calculated based on the preceding national census output areas. Each participant is assigned a score corresponding to the output area in which their postcode is located. A higher index indicating a greater level of deprivation (**Supplementary Information**).

Venous blood samples were taken and stored for subsequent assays, including genome-wide genotyping.[21] Overall, 488,377 (97%) participants were successfully genotyped: 49,979 using the UK BiLEVE chip and 438,398 using the UKB axiom chip. Pre-imputation quality control, phasing, and imputation of the UKB genetic data have been described.[22] Detailed information on the full measurements can be found at UKB showcase.[23]

We considered 43 baseline anthropometric and metabolic traits including BMI, waist circumference (WC), hip circumference (HC), diastolic blood pressure (DBP), systolic blood pressure (SBP), whole body fat mass (WBFM), whole body fat-free mass (WBFFM), trunk fat mass (TFM), trunk fat-free mass (TFFM), mean grip strength (mean of left and right grip strength) basal metabolic rate (BMR), forced expiratory volume in 1-second (FEV1), forced vital capacity (FVC), peak expiratory flow (PEF), low-density lipoproteins cholesterol (LDL), high-density lipoproteins cholesterol (HDL), total cholesterol (TC), total triglycerides (TG), apolipoprotein A (Apo A), apolipoprotein B (Apo B), Lipoprotein A (Lipo A), sex hormone binding globulin (SHBG), testosterone, oestradiol, insulin-link growth factor 1 (IGF-1), alanine aminotransferase (ALT), alkaline phosphatase (ALP), aspartate aminotransferase (AST), gamma glutamyltransferase (GGT), albumin, total bilirubin, direct bilirubin, total protein, urate, urea, creatinine, cystatin C, phosphate, rheumatoid factor, C-reactive protein (CRP), Vitamin D, calcium as diabetes risk factors. Detailed information of the above traits and how they were measured would be referred to the UKB showcase.[23] The Data Fields of above-mentioned traits were shown in **Supplementary Information**.

The UK Biobank Pharma Proteomics Project (UKB-PPP) is a precompetitive consortium of 13 biopharmaceutical companies funding the generation of blood-based proteomic data from UK Biobank volunteer samples,[10] which included 54,219 UK Biobank participants and 2,923 unique proteins measured by antibody-based Olink Explore 3072 PEA. Additional information (e.g., participant selection, Olink proteomics assay, and quality control) was described elsewhere.[10] Each protein level was inverse-rank normalized in normalized protein expression unit (NPX). We focused on 33,301 participants in the UKB-PPP who are 1) European ancestry (defined by the first 15 principal components, where the method was described[24] to calculate a 15-coordinate median of the self-reported White and 1000G EUR samples); 2) with the Olink Batch PlateID from one to six (participants from the pilot study, Batch PlateID = 0, and from the Covid-19 imaging sample, Batch PlateID = 7 were excluded)[25]; 3) No spurious sample by deCODE; 4) no batch from cherry-picked by the UKBB-PPP consortium and no Covid imaging sample; 5) the randomly selected sample; 6) passing the Novo Nordisk in house sample and genetic QC procedures. All participants provided informed consent.

Hemoglobin A1C (HbA1c) was measured in red blood cells by HPLC on a Bio-Rad VARIANT II Turbo analyzer and glucose was assayed in serum by hexokinase analysis on a Beckman Coulter AU5800.[21] Samples were assumed to be non-fasting, because participants were not advised to fast before attending. They were asked to record the last time they ate or drank anything other than water before attending the clinic and those answers were used to derive ‘fasting time’ prior to sampling. During routine quality control checks, the UKB laboratory team observed that, due to a sample processing error, ∼ 8% of the glucose assay results were lower than expected and therefore a ‘dilution correction factor’ was provided and applied to these results.[26] The HbA1c samples were not affected.

Baseline T2D prevalence (pT2D) was considered of those who were 1) baseline reported doctor-diagnosed diabetes: 2) whose date of first reported diabetes (ICD-10, E11: non-insulin-dependent diabetes mellitus) ahead of the date of attending assessment; 3) random blood glucose >= 11.1 mmol/L OR HbA1c >= 48 mmol/mol. It is possible to misclassify a few type 1 diabetes prevalence cases into pT2D. However, no self-reported diabetes cases were approximately diagnosed before 25 years old in the UKBPPP subsample. T2D incidence (iT2D) cases were identified in according the ICD-10 code (E11: non-insulin-dependent diabetes mellitus).[27] The censoring/leaving time was considered to be earliest among the following dates: 1) T2D diagnosed date; 2) censoring date from the hospital inpatient data; 3) death date; 4) lost to follow-up date. Details of how to generate the censoring time is shown in **Supplementary Information**.

#### The China Kadoorie Biobank

The baseline survey of China Kadoorie Biobank (CKB) was established during 2004 - 2008 in ten geographically diverse regions in China (five were urban regions and five were rural regions). 512,715 participants aged 30–79 years were recruited. Sociodemographic characteristics (e.g., education, income), lifestyle (e.g., smoking, alcohol consumption, physical activity), and personal and family medical history (e.g., family history of diabetes), self-reported health status, as well as physical and blood measurements (e.g., BMI, and random blood glucose, [HbA1c was not available]) were measured in the baseline survey. Prior to starting the project, central ethics approvals were obtained from Oxford University and the China National Center for Disease Control and Prevention (CDC). In addition, approvals were also obtained from institutional research boards at the local CDCs in the ten regions. All participants provided written informed consent.[28, 29] The detailed information of the above mentioned characteristics are available in **Supplementary Information** and the CKB data showcase.[30]

105,408 participants were genotyped used custom-designed arrays on the Affymetrix (now Thermo Fisher Scientific) Axiom platform, with content selection based on similar overall principles to those used for the UK Biobank array design but adapted to optimize performance for individuals of East Asian ancestry. The genotyped sample was from two array version (array version 1 was a nested case-control design of cardiovascular disease and chronic obstructive pulmonary disease; array version 2 contained 72,000 samples; in total there are ∼ 100,000 unique samples after quality control with ∼ 74,000 randomly selected). Detailed information was reported.[28, 29]

The Olink proteomics assay was conducted among 3,977 CKB participants from a further nested case-control sub-cohort study of myocardial infarction (1,951 incident cases of ischemic heart disease and 2,026 control, all with genotyping and had no prior history of cardiovascular diseases, no use of lipid-lowering drugs (e.g. statins) at time of sample collection).[29] In this study, to account for the potential risk of selection bias, we focused on 2,029 randomly selected participants. The protein data was flagged as having a quality control warning if the incubation control deviates more than a predetermined value (±0.3) from the median value of all samples on the plate. The pre-processed data were provided in the arbitrary unit normalized protein expression (NPX) on a log2 scale. Protein measures were analysed as rank inverse normal transformed residuals following linear regression on sex, age, age square, and region (10-level categorical variable). In this study, 2,921 unique proteins data were available (ERVV-1, HLA-A were not available in CKB).

T2D prevalence (pT2D) was obtained via self-reported diabetes data (i.e., participant has history of diabetes (reported OR random blood glucose >= 11.1 mmol/L OR fasting blood glucose >= 7.0 mmol/L)) in the baseline. As above, we cannot rule out misclassification of type 1 diabetes prevalence into as pT2D, though no self-reported diabetes cases were approximately diagnosed before 25 years old in the CKB Olink subsample. We identified T2D incidence (iT2D) cases based on the ICD10 (E11) code (follow-up till 31 Dec 2018). The following information would be considered as censored/leaving whenever the date of death, the date of loss to follow-up, and the date of T2D diagnosed by 31 Dec 2018.

#### Genetic variants associated with proteins, diabetes traits, and metabolic/anthropometric traits

##### Proteome

In the UKB-EUR, we obtained the cis-pQTLs (protein quantitative trait loci) from genome-wide association studies (GWAS) of 48,645 EUR participants with 2,923 unique proteins measured. The protein level was inverse-rank normalized, including NPX data below the limit of detection. GWAS analyses were performed using REGENIE v.3.2.4.[31] The model included the following covariates: age, age^2^, ageD×Dsex, genotype array batch, and the first 10 genetic principal components. We restricted the analysis to the variants: MAFD(minor allele frequency) >D0.5%, Hardy–Weinberg equilibrium test PD>D10E−12, <5% missingness and linkage-disequilibrium (LD) pruning (1,000 variant windows, 100 sliding windows and r2D<D0.8). We identified approximately independent cis-pQTLs by using LD clumping to select instrumental variables (IVs) for Mendelian randomization analyses. We restricted the IVs to the cis-region only, using a +/-500kb window of the protein-coding gene, and performed LD clumping with the following criteria: MAF > 0.001, R2 < 0.01, and p-value < 5e-08 using PLINK v1.9.[32] The LD reference is a 10k-sample of European origin, randomly selected from the UK Biobank (Novo Nordisk in-house LD reference). The GWAS summary statistics have not yet been published but scientifically applied as Novo Nordisk in-house data.

In the CKB-EAS, we extracted the cis-pQTLs from the GWAS study with ∼4,000 adults.[33] Genome-wide association analyses were performed for the rank inverse normal transformed protein levels with 11 principle components and genotyping array version as covariates (other covariates were adjusted for in the transformation of protein measures described above). Post GWAS filtering removed single nucleotide polymorphisms (SNPs) with effective minor allele count <20 (MAF × INFO score × 2 × N <20). pQTL loci were identified by LD clumping around association signals reaching genome-wide significance. Clumping was done using internal CKB LD reference of 72K unrelated individuals with the use of PLINK v1.9.[32] Sentinel variants in linkage disequilibrium (LD R^2^>0.05, P-value >0.05) within 5Mbp were identified; for major histocompatibility complex (MHC) proteins 20Mb was used. Association signals were categorised as cis-pQTLs if the sentinel variant was within 500Kbp of the structural gene for the assayed protein. Detailed has been previously described.[33]

##### Diabetes traits

Eight diabetes traits (T2D,[34] HbA1c,[35] fasting glucose,[35] fasting insulin (BMI adjusted, BMI unadjusted is not available),[35] random glucose (BMI adjusted, BMI unadjusted is not available),[36] proinsulin,[37] modified Stumvoll insulin sensitivity index (ISI)[38] and the fold-change in insulin concentration (IFC)[38]) GWAS were available for the EUR population. For ESA population GWAS, four diabetes traits (T2D,[34] HbA1c,[39] fasting glucose,[39] fasting insulin (BMI adjusted, BMI unadjusted is not available[35]) were available. These GWAS were obtained from the MAGIC (the Meta-Analyses of Glucose and Insulin-related traits Consortium), DIAGRAM (DIAbetes Genetics Replication And Meta-analysis) consortium, Biobank Bank Japan, Taiwan Biobank (**Supplementary Table 1**).

##### Metabolic/anthropometric traits

For EUR, the GWAS summary statistics of 28 cluster leading (the unique anthropometric/metabolic traits in top weight with accumulated weights ≥50%) metabolic/anthropometric traits were available. The summary statistics of BMI and WHR, which were obtained from a meta-analysis combining the results from the Genetic Investigation of ANthropometric Traits (GIANT) consortium and UKB.[40] The summary of others metabolic/anthropometric traits (WC, HC, WBFM. WBFFM, TFM. TFFM, BMR, pulse rate, TG, TC, LDL, HDL, Apo A, Apo B, ALT, ALP, AST, GGT, cystatin C, creatinine, urea, urate, SHBG, calcium, albumin, and total protein) were downloaded from Pan-UK Biobank,[41] which releases the results of a large genome-wide analysis for a wide range of outcomes in a diverse group (e.g., different ancestries) of individuals from the UK.

For EAS, the GWAS summary statistics of 17 cluster-specific metabolic/anthropometric traits (BMI, WHR, WC, HC, TG, LDL, HDL, TC, ALT, AST, ALP, GGT, creatinine, urea, calcium, albumin, total protein) were available. Two additional metabolic/anthropometric traits (body fat rate (BFR) and resting heart rate (RHR)) were applied, which were measuring similar characteristics as WBFM and pulse rate did. The summary statistics of BMI, TG, LDL, HDL, TC, ALT, AST, GGT, creatinine, urea, and albumin were from the largest EAS GWAS combing the Taiwan Biobank and Biobank Japan.[39] For WHR, WC, HC, BFR, and RHR, the summary statistics were available in the Taiwan Biobank cohort.[39] Lastly, the summary statistics of ALP, calcium, and total proteins were from the Biobank Japan.[42]

Details (e.g., sample size, unit) of these GWAS data were available in the **Supplementary Information** and **Supplementary Table 1**. Detailed description of specific studies’ design and characteristics of studies’ participants were provided in the original publications.

### Statistical analysis

#### Multivariable-adjusted regression

We applied multivariable-adjusted regression in cross-sectional (linear and logistic regressions) and prospective cohort (cox regression) designs to identify proteins associated with diabetes as well as glycaemic traits. In UKB-EUR, baseline HbA1c (mmol/mol), random glucose (mmol/L), and self-reported pT2D were assessed outcomes in cross-sectional design; iT2D was the assessed outcome in prospective cohort design. We considered different levels of adjustments via applying two models. In Model 1, we adjusted for baseline age, age square, sex, assessment centre, fasting time, dilution factor, family history of diabetes. In Model 2, we additionally adjusted for education (ISCED), Townsend deprivation index, smoking, alcohol consumption, summed MET time, self-reported health rating. We considered the statistical cut-off with FDR and Bonferroni correction with the number of proteins tested (n = 2,923). We picked up the outcome specific proteins passing the Bonferroni correction in both models into the clustering step.

In the CKB-EAS, similar analyses were conducted though HbA1c was not available. In Model 1 baseline age, age square, sex, study region, ambient temperature, fasting time, family history of diabetes, and plate ID were adjusted for. In Model 2, highest education, household income, smoking, alcohol consumption, physical activity (MET time), and self-reported health status were additionally adjusted for. Given the small sample size (n = 2,029), the numbers of pT2D (n = 130) and iT2D (n = 65) were not large enough to account for the variation caused by plate ID (114 plate IDs across eight panels). Therefore, we utilized the residual to account for the variation, where we obtained the residual of the protein levels via regressing on plate ID, study region, temperature, and fasting time; we then regressed the outcome (dependent variable, e.g., pT2D) on the residual, as obtained in above, and the model specific covariates. We considered the statistical cut-off with FDR and Bonferroni correction with the number of proteins tested (n = 2,921).

We acknowledged confounded estimates were likely captured due to BMI, though, in some cases, BMI plays as a mediator adjusting for BMI would introduce collider bias.[43] There is no foolproof model to account for all. We maximized the number of proteins obtained from the observational designs without adjusting for BMI.

#### Bayesian non-negative matrix factorisation (bNMF)

The nonnegative matrix factorisation (NMF) is a soft clustering method (i.e., a feature can be assigned to more than one clusters), which decomposes a high-dimensional data matrix into two lower-dimensional matrices with non-negative elements (i.e., V ≈ W × H; V, W, and H are nonnegative matrixes; for example, W was a matrix of clusters *vs* proteins, H was a matrix of clusters *vs* metabolic/anthropometric traits, which were filled by the contributions of each protein/trait in the corresponding cluster). Various methods (e.g., Frobenius norm, the Kullback-Leibler divergence) can be applied to minimize the error i.e., Min |IV - WH|Iloss, where the number of the clusters would be obtained. The Bayesian non-negative matrix factorisation (bNMF) was designed to obtain an optimal number of clusters, *K* that best explain V at the balance between an error measure, |IV - WH|I^2^ (i.e., iteratively regresses out irrelevant components in representing V with an automatic relevance determination technique, which enables an optimal inference for the number of clusters *K*, and a penalty for model complexity derived from a nonnegative half-normal prior for W and H.) The pipeline and statistical details can be found elsewhere.[6, 7]

In this study, the bNMF clustering was conducted in UKB-EUR but not CKB-EAS due to lack of metabolic traits in the CKB-EAS participants with proteins measurement. In the main analyse, we generated the non-negative matrix V based on the associations of all diabetes-associated proteins (n = 1,793, i.e., the protein associated (p-value < 1.7 × 10^^-5^, Bonferroni-corrected) with at least one diabetes traits (i.e., HbA1c, random glucose, pT2D, iT2D) with 43 metabolic/anthropometric traits (BMI, WC, HC, WBFM, WBFFM, TFM, TFFM, mean grip strength, BMR, PR, SBP, DBP, LDL, HDL, TC, TG, Apo A, Apo B, Lipo A, SHBG, testosterone, oestradiol, IGF-1, ALT, ALP, AST, GGT, albumin, total bilirubin, direct bilirubin, total protein, urate, urea, creatinine, cystatin C, phosphate, rheumatoid, CRP, vitamin D, calcium, FEV-1, FVC, PEF). The protein – trait association was from a linear regression adjusted for the same covariates as applied in Model 2 in the observational analysis, where the corresponding standardized z-scores (i.e., beta in SD unit / se in SD unit) were fit into the matrix V*. In order to guarantee matrix V non-negative, the number of metabolic/anthropometric traits were doubled (i.e., V was a 1,793 by 43×2 matrix) by concatenating two separate modifications of the original matrix V*: one containing all positive standardized z-scores (zero otherwise) and the other all negative standardized z-scores multiplied by −1 (i.e., all the metabolic/anthropometric traits should be two features e.g., BMI and Reduced-BMI). We applied 100 iterations in bNMF and considered the top 10% proteins in terms of ranking in each cluster of matrix W as the most identical cluster-leading proteins and those in top weight with accumulated weights more than 50% in matrix H as the cluster-leading dominant/representative traits.

In additional analyses, we accounted for trait – trait and protein – protein correlations, where the highly correlated (The square of Pearson correlation coefficients >= 0.80 with age, sex, and assessment centre adjusted) traits (**Supplementary Table 2**) and proteins (**Supplementary Table 3**) were excluded in the bNMF clustering analyses. Besides, matrixes of four diabetes traits (e.g., HbA1c, random glucose, pT2D, iT2D) associated proteins (n = 1,466, 708, 787, 815 respectively) (**Supplementary Table 4**) with 43 metabolic/anthropometric traits were generated and then bNMF clustering was conducted. The corresponding leading traits and proteins in each model were shown in **Supplementary Table 5** and **6** respectively. The number of overlapping leading proteins across all bNMF models (e.g., HbA1c vs glucose outcomes; with and without protein–protein and trait–trait correlations) were shown in **Supplementary Fig. 1**. All the proteins, the top 10% precent, from the main model (i.e., 1,793 proteins *vs* 43 metabolic/anthropometric traits) and the additional (diabetes traits (e.g., HbA1c) associated proteins (1,466) *vs* 43 metabolic/anthropometric traits) analyses were progressed to the colocalization step.

#### Colocalization

We checked whether the above-obtained diabetes associated proteins share a causal variant with the diabetes traits as well as the cluster-leading metabolic/anthropometric traits in a genomic region using the hypothesis prioritisation for multi-trait colocalization (HyPrColoc).[44] HyPrColoc is a Bayesian method for identifying shared genetic associations between complex traits in a particular gene region using summary GWAS results. Statistical detail was described elsewhere.[44]

The HyPrColoc method presents two major posterior probability statistics to interpret the evidence of colocalization: 1) regional probability (P_R_), it allows more than one causal variant shared by all traits (in our case, protein and trait), which is due to the two or more traits have distinct causal variants in strong LD with one another; 2) alignment probability (P_A_), it is a safeguard that quantifies the probability of alignment at a single causal variant between the shared association. The posterior probability of full colocalization (PPFC) would be calculated via PPFC = P_R_ x P_A_. There are two strategies to set up the prior probability, nonuniform prior probability and uniform prior probability. The nonuniform prior probability assumes that prior probability to not vary by genetic variant, nor by the specific collection of colocalized traits of a given size, but by the number of colocalized traits (i.e., a SNP associated with a total of k traits has a prior probability that depends on the number k but not the specific collection of traits). The uniform prior does not set variant-level information and implicitly account for large differences in the causal configuration space between hypotheses, which limits the loss in power of the PPFC for very large traits of testing. The uniform estimate has a higher risk of type 1 error (false positive) of colocalization.[44] As such, we only considered using the nonuniform prior probability statistics to check the evidence of genetic alignment. The pairwise colocalization (one protein *vs* one trait) inflates the false negative rate[44] but reduces the risk of the missing shared genetic variants across multiple traits of different GWAS. We did not apply the posterior probability of full colocalization (PPFC) (PPFC = P_R_ x P_A_) to pick up the colocalized proteins. In practices, we found extreme cases that PPFC cannot capture colocalized pairs. For example, the UKB-PPP measured SHBG protein with SHBG protein measured in the whole UKB sample, as well as PCSK9 protein with LDL, in both cases, P_A_ are equal to 0 and P_R_ are equal to 1. Merely rely on PPFC would lose candidate pairs. The nonuniform P_A_ shares conceptual similarities with the posterior probability of H4 (Coloc).[45] Both focus on the likelihood of shared genetic influence or causal variants between traits based on alignment and signal similarity.

We introduced a colocalization tier (CT) system based on the confidence of genetic alignment to pick up the candidate colocalized proteins. Colocalization Tier 1 (CT1): nonuniform P_A_ >= 0.7 AND nonuniform P_R_ >= 0.7; Colocalization Tier 2 (CT2): nonuniform P_A_ >= 0.7 OR nonuniform P_R_ >= 0.7 (mainly pick-up the cases with nonuniform P_R_ >= 0.7 only; those with nonuniform P_A_ >= 0.7 were expected to have nonuniform P_R_ >= 0.7, which would likely be classified into CT1); We called “Relaxed Colocalization” when the probability statistics fulfilling CT1 or CT2; we called “Strict Colocalization” when the probability statistics fulfilling CT1.

We processed two steps of colocalization. Step 1 was conducted on the cluster-leading proteins (first 10% in terms of ranking) with diabetes traits. Step 2 focused on the proteins, passing “Relaxed Colocalization” with at least one of the diabetes traits, and colocalization analyses were conducted with cluster-leading metabolic/anthropometric traits. The two steps colocalization analyses would also be applied in CKB-EAS population, where the genetic associations of the proteins were obtained from the CKB-EAS. In EAS population, four diabetes traits GWAS and 19 metabolic/anthropometric traits GWAS were available. We relaxed the cut-off in the colocalization tiers from 0.70 to 0.65, considering the smaller sample size of the EAS GWAS. Last but not the least, no colocalization analysis was conducted on those proteins – trait pairs whenever the gene region of the protein was in the X chromone given the majority GWAS data had no summary statistics of the X chromone (**Supplementary Information**).

#### Mendelian randomization

The instrument strength was assessed by the mean F-statistic, which was equal to the mean of the individual F-statistic of each cis-pQTL(protein quantitative trait loci) (individual F statistic was equal to cis-pQTL specific estimate: 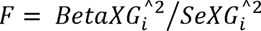. The variance explained by genetic instrument (R^2^) was calculated via *R*^2^ = *K* * *F* ⁄ (*N* - *K* - 1 + *K* * *F*) (*F* is the mean F-statistic in above, *K* is equal to the number of cis-pQTLs applied, *N* is the GWAS sample size). An F-statistic < 10 indicates risks of weak instrument bias,[46] though this threshold is arbitrary. In general, the higher the R^2^ and F-statistic the less likelihood of weak instrument bias. The mean F-statistics of all the cis-pQTLs of the colocalized proteins 53 (HHEX has no available cis-pQTL) and 9 (FST has no available cis-pQLT) colocalized proteins in UKB-EUR and CKB-EAS were large than 10 (**Supplementary Information** and **Supplementary Table 7, 8**).

Two-sample Mendelian randomization (2SMR) analysis was conducted to assess the effect of colocalized proteins with all the diabetes traits and cluster-leading metabolic/anthropometric traits. For sample 1, we identified the independent (r^2^ <= 0.01) and genome-wide significant (p < 5 x 10^-8^) *cis*-protein quantitative loci (cis-pQTLs) derived from the GWAS in UKB-PPP (for EUR) and in CKB (for EAS) as the genetic instruments for proteins. For sample 2, the summary statistics of these cis-pQTLs were obtained from the GWAS of the corresponding traits (**Supplementary Information** and **Supplementary Table 1**). For proteins with <= 2 cis-pQTLs, we applied inverse-variance weighted (IVW) regression of the Wald ration (WR) for each cis-pQTL under a multiplicative random-effects model[47] to obtain the causal estimates. No pleiotropy robust analysis was available. For proteins with three independent cis-pQTLs, MR-Egger[48] was added to account for the risk of pleiotropy. When those proteins with > 3 cis-pQTLs, Mendelian randomization pleiotropy residual sum and outlier (MR-PRESSO)[49] were added. Whenever the MR-PRESSO global test indicated a risk of horizontal pleiotropy (i.e., the effect of the cis-pQTL on the outcome of interest is mediated via others apart from the protein of interest), the outliers (i.e., cis-pQTLs) would be excluded and the corresponding IVW, MR-Egger, and MR-PRESSO analyses were re-ran. Considering the massive computing burden, we did not identify the proxy of 11 colocalized robust proteins that the corresponding cis-pQTLs cannot be identified in some specific GWAS (**Supplementary Table 9**). Thus, the corresponding estimates were missing. In the CKB proteomic GWAS, all the proteins tested were with one cis-pQTL and WR was applied. Whenever the cis-pQTL was not available in the outcome GWAS, a proxy with R^2^ > 0.8 would be applied (**Supplementary Table 10** and **Supplementary Information**).

In addition, bidirectional/reverse 2SMR was considered to explore potential inverse effects (i.e., traits to proteins). The independent and genome-wide significant SNPs of the traits were extracted from the corresponding GWAS studies (**Supplementary Information**), where the SNP – exposure (trait) (sample 1) associations were obtained. The SNP – outcome (protein) (sample 2) associations would be extracted in the UKB-PPP and CKB proteomics[33] GWAS. IVW, weighted median (WM),[50] and MR-Egger (nSNP >= 3) would be applied. MR-PRESSO was not considered given the large number of genetic instruments in most of the exposures, where extensive computing load was needed.

In order to pick up the candidate associations among the large number of tested protein – trait pairs, we developed a four-levels Tier system (MRT1 – MRT4) incorporating all MR estimates including the main (IVW) and sensitivity (MR-Egger, WM, MR-PRESSO) analyses results, as well as multiple testing (Bonferroni-corrected). Details of the Tier system was described in the **Supplementary Information** and **Supplementary Table 11**. For Bonferroni-corrected p values, in UKB-EUR, we considered the p value < 2.6 × 10^^-5^ (i.e., 0.05 / (53 × 36)) (HHEX has no available cis-pQTL) as statistically significant when accessing the effect of protein to trait; we considered the p value < 2.6 × 10^^-5^ (i.e., 0.05 / (54 × 36)) statistically significant when assessing the effects of traits on proteins. In CKB-EAS, we considered the p value < 2.2 × 10^^-4^ (i.e., 0.05 / (9 × 23)) (FST has no available cis-pQTL) as statistically significant when accessing the effect of protein to trait; we considered the p value < 2.2 × 10^^-4^ (i.e., 0.05 / (10 × 23)) statistically significant when assessing the effects of traits on proteins. The associations classified into MRT1 and MRT2 were considered stronger causal evidence, which supported by the main analysis as well as by the pleiotropy robust sensitivity analyses whenever assessable. Whenever the protein – trait pair estimates were with evidence of outliers identified by MR-PRESSO, the corresponding tier was based on the outlier-corrected estimates. MR Steiger[51] test would be applied to check the directionality whenever diabetes associated bidirectional pairs (e.g., protein – T2D) were identified.

## Results

### Baseline characteristics

Overall, 33,301 participants (54.2% female, mean age 56.8 (SD 8.0) years, mean BMI 27.3 (SD 4.7) kg/m^2^) with plasma proteins (Olink Explore 3072 PEA) measured by the UK Biobank Pharma Proteomics Project (UKB-PPP) were included in the analyses; 1,803 (5.4%) had self-reported prevalent T2D (pT2D) at baseline, and 1,082 developed new onset, incident, T2D (iT2D) during follow-up. Among the 2,029 CKB participants (62.1% female, mean age 51.3 (10.5) years, mean BMI 23.8 (3.4) kg/m^2^) with plasma proteins measured using the same Olink Explore panel, 130 (6.4%) participants had self-reported pT2D and 65 developed iT2D during follow-up. **Table 1** provides further baseline characteristics of the proteomics-assayed participants in both populations.

**Table 1.** Baseline characteristics of study participants with proteomic measurements in the UK Biobank and China Kadoorie Biobank.

| UK Biobank (UKB-EUR) |  | China Kadoorie Biobank (CKB-EAS) |  |
| --- | --- | --- | --- |
| Covariates | Mean (SD) / (%) | Covariates | Mean (SD) / (%) |
| N | 33,301 | N | 2,029 |
| Age | 57 (8) | Age | 51 (10) |
| Female | 54.2 | Female | 62.1 |
| BMI | 27.3 (4.7) | BMI | 23.8 (3.4) |
| Smoking |  | Smoking |  |
| Never | 53.6 | Never smoker | 64.2 |
| Previous | 35.5 | Occasional smoker | 5.4 |
| Current | 10.5 | Ex-regular smoker | 5.1 |
| Prefer not to answer / Missing | 0.4 | Regular smoker | 25.3 |
| Alcohol consumption |  | Alcohol consumption |  |
| Never | 6.5 | Never regular | 46.2 |
| Special occasions only | 10.5 | Ex-regular | 1.7 |
| 1 to 3 times a month | 10.9 | Occasional | 32.0 |
| 1 or 2 times a week | 26.3 | Monthly | 3.0 |
| 3 or 4 times a week | 23.8 | Reduced intake | 1.8 |
| Daily or almost daily | 21.7 | Weekly | 15.2 |
| Prefer not to answer / Missing | 0.1 |  |  |
| Physical activity (hours /week) | 13.8 to 59.6 | Physical activity (metabolic equivalent of task, hours /day, IQR) | 10.1 to 30.3 |
| Education |  | Education |  |
| None of below | 16.7 | No formal school | 16.4 |
| O levels / GCSEs or equivalent / CSEs or equivalent | 16.7 | Primary school | 31.4 |
| A levels/AS levels or equivalent | 5.6 | Secondary school | 29.3 |
| Other professional qualification eg:<br>nursing, teaching | 12.1 | High school | 14.9 |
| College or University degree / NVQ or<br>HND or HNC or equivalent | 47.8 | Technical school / college | 4.3 |
| Prefer not to answer / Missing | 1.0 | University | 3.5 |
| Family history of diabetes |  | Family history of diabetes (Yes) | 153 |
| At least 1 member has diabetes | 20.0 |  |  |
| At least 1 member has no diabetes but<br>other diseases\$ | 72.1 | | |
| Do not know / Not to answer / None<br>above / Missing | 7.9 |  |  |
| Baseline T2D (number and %) | 1,545 / 4.7 | Baseline diabetes (number and %) | 63 / 3.1 |
| Screening-detected baseline T2D* (number<br>and %) (pT2D) | 1,803 / 5.4 | Screening-detected baseline diabetes*<br>(number and %) | 130 / 6.4 |
| Incidence case of type 2 diabetes (number)<br>(iT2D)* | 1,082 | Incidence case of type 2 diabetes<br>(number) | 65 |
| Random glucose, mmol/l | 5.1 (1.2) | Random glucose mmol/l | 6.0 (2.3) |
| Random glucose in participants with<br>baseline diabetes mmol/l | 7.6 (3.4) | Random glucose in participants with<br>baseline diabetes mmol/l | 11.7 (5.4) |
| HbA1c mmol/mol | 36.0 (6.6) |  |  |
| HbA1c in participants with baseline diabetes<br>mmol/mol | 53.2 (15.3) |  |  |

\* In UKB: Self-reported diagnosed T2D + First reported T2D (ICD-10 E11) before recruitment + screening detected diabetes (random blood glucose $\geq 11.1$ mmol/l OR HbA1c $\geq 48$ mmol/mol; in CKB: Participant has history of diabetes (self-reported OR random blood glucose $\geq 11.1$ mmol/L OR fasting blood glucose $\geq 7.0$ mmol/L). T2D prevalence (pT2D) as the analyses applied in the manuscript. T2D incidence (iT2D) as the analyses applied in the manuscript.
\$ Other diseases included Heart disease, Stroke, High blood pressure, Chronic bronchitis/emphysema, Alzheimer's disease/dementia, Parkinson's disease, Severe Depression, Lung cancer, Bowel cancer, Breast cancer.

### UK Biobank – EUR

To identify candidate proteins associated with T2D and/or glycaemic traits, observational analyses were adopted (**Methods**). Overall, 1466, 708, 787, 815 proteins were associated (Bonferroni-corrected p-value<1.7 × 10^-5^ threshold) with HbA1c, random glucose, pT2D, and iT2D respectively, with 1,793 unique proteins associated with at least one and 315 were associated with all four outcomes (**Supplementary Fig. 2** and **Supplementary Table 4**). Detailed observational estimates are in **Supplementary Table 12**.

For the 1,793 proteins associated with at least one phenotype (HbA1c, random glucose, pT2D or iT2D), we tested their association with 43 anthropometric and metabolic traits. Next, we used this information to cluster the proteins according to their phenotypic associations to assess the likely roles of these 43 anthropometric and metabolic traits in explaining the observed protein-diabetes associations (**Methods**). Overall, we identified five clusters and used the anthropometric/metabolic traits that collectively accounted for 50% of the cluster weights to label these as “Adiposity”, “Reduced-adiposity”, “Lipids”, “Liver”, and “Kidney” clusters (**Fig. 2**). All 1,793 proteins were represented in each cluster with different weights, however, the top ∼180 proteins (top 10%) by importance ranking for each cluster were considered representative of that cluster. These 180 proteins accounted for 33.3% of the weights in the adiposity cluster (where 72 were specific to the adiposity), while 81.1% (161 cluster specific), 24.8% (157 cluster specific), 49.0% (118 cluster specific), and 43.1% (117 cluster specific) of the weights in the Reduced-adiposity, Lipids, Liver, and Kidney clusters, respectively (**Fig. 2**, **Supplementary Table 5** and **6**). Overall, we extracted 906 unique proteins (**Supplementary Table 6**) across five models in the main and additional analyses (i.e., besides the main model we additionally included all the unique proteins across other models tested), which were taken forward into the genetic analyses, using their genetic features identified from GWAS (**Methods**). Among the 906 proteins, 21 were encoded on the X chromosome and were omitted due to lack of available genetic information (**Supplementary Information**), leaving 885 proteins for further analyses.

**Fig. 2.**
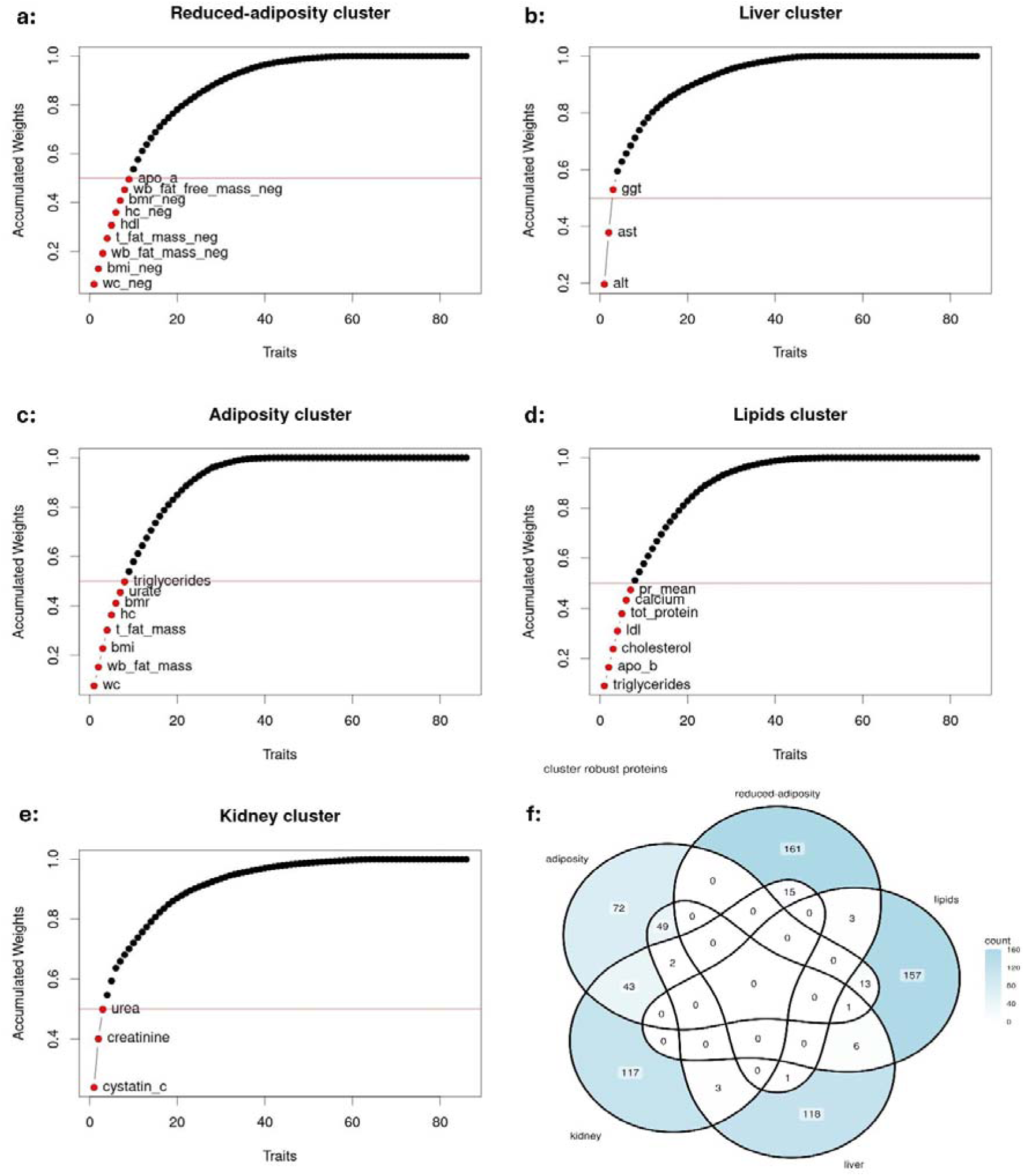
Leading metabolic/anthropometric traits (accumulated weights ≥50%) in five main clusters (a-e) and the number of overlapping leading proteins across five clusters in the main model (f) The main model that all proteins (n = 1,793) associated with at least one of the diabetes traits (HbA1c, glucose, pT2D, iT2D) in the UK Biobank were used to generate the matrix V for the bNMF clustering. “_neg”: negative traits, waist circumference (wc), whole body fat mass (wb_fat_mass), whole body fat free mass (wb_fat_free_mass), body mass index (bmi), tunk fat mass (t_fat_mass), hit circumference (hc), low-density lipoproteins (ldl), high-density lipoprotein (hdl), basal metabolic rate (bmr), hip circumference (hc), apolipoprotein A (apo_a), apolipoprotein B (apo_b), sex hormone binding globulin (shbg), mean of pulse rate (pr), mean of diastolic blood pressure (dbp_mean), mean of systolic blood pressure (sbp_mean), total protein (tot_protein), direct bilirubin (dir_bilirubin), total bilirubin (tot_bilirubin), alanine aminotransferase (alt), aspartate aminotransferase (ast), alkaline phosphatase (alp), gamma-glutamyltransferase (ggt), cystatin C (cystatinc)

To identify which protein–trait pairs shared the same causal genetic variants, we conducted two-steps colocalization using HyPrColoc with nonuniform priors.[44] We developed Colocalization Tiers (CT) to pick up the candidate protein-trait pairs with evidence of genetic alignment with ≥1 diabetes trait(s). Colocalization Tier 1 (CT1) required regional probability (PR) ≥0.7 AND alignment probability (PA) ≥0.7, while Colocalization Tier 2 (CT2) required PR ≥0.7 OR PA ≥0.7 (A relaxed threshold of 0.65 was applied in CKB-EAS considering the smaller CKB protein-GWAS study). In the first step, we tested the 885 cluster-leading proteins for genetic colocalization with eight diabetes related traits (HbA1c, fasting glucose, random glucose, fasting insulin, proinsulin, modified Stumvoll insulin sensitivity index (ISI), the fold-change in insulin concentration (IFC), and T2D) in EUR population. Overall, 54 proteins passed the “Relaxed Colocalization” criteria (i.e., fulfilling CT1 or CT2), among which 12 proteins passed the “Strict Colocalization” criteria (i.e., fulfilling CT1). In the second step, to identify specific protein-cluster phenotype pairs that shared the same causal genetic variants, we took the 54 proteins into the colocalization analysis with the 28 cluster-leading metabolic/anthropometric traits (i.e., the unique anthropometric/metabolic traits in a given cluster that had accumulated weights ≥50%). Overall, for the cluster leading metabolic/anthropometric traits, 38 and 12 proteins passed the “Relaxed Colocalization” and “Strict Colocalization” respectively, resulting in five (i.e., GRP, INHBC, MEGF9, NELL1, SHBG) passing the “Strict Colocation” from both steps (**Supplementary Table 13**). The detailed colocalization results are provided in **Online Results**.

For the 54 proteins with (relaxed) colocalization evidence, we conducted bidirectional MR to investigate three-way relationships (i.e., protein – metabolic/anthropometric traits – diabetes traits) (**Methods**). We developed MR Tiers to pick up the candidate protein – trait pairs with potential causal relationships. MR Tier 1 (MRT1) indicates strong MR evidence accounting for multiple testing (UKB-EUR: Bonferroni-corrected p value < 2.6 × 10^-5^; CKB-EAS Bonferroni-corrected p value < 2.2 × 10^-4^) across all applied MR methods; MR Tier 2 (MRT2) includes all protein-trait pairs apart from pairs in MRT1 whose raw (i.e. not corrected for multiple testing) MR p-values were below 0.05 (**Methods** and **Supplementary Information**). We tested whether 53 genetically predicted proteins (HHEX has no available *cis*-pQTLs) were associated with 36 traits (8 diabetes traits and 28 cluster-leading traits), and found 76 MRT1 and 359 MRT2 protein – trait pairs.

Incorporating colocalization evidence, nine pairs were in MRT1 and CT1, including three proteins (CD34, KCTD5, SCT) associated with T2D (**Supplementary Fig. 3**); and 157 pairs were in MRT1/MRT2 and CT1/CT2 (**Supplementary Table 14**). We next assessed the effects of traits on proteins using reverse MR and found 101 MRT1 and 485 MRT2 trait – protein pairs. Supported by colocalization evidence, three pairs were in MRT1 and CT1 (no T2D – protein pair); 70 pairs were in MRT1/MRT2 and CT1/CT2 (**Supplementary Table 14**). In sum, 41 candidate proteins were genetically associated with T2D and/or glycaemic traits (**Supplementary Table 15**). All bidirectional effects associated with T2D were supported by the MR Steiger directionality test (**Supplementary Table 16**). Statistical details of MR estimates are shown in **Online Results**.

We triangulated observational and genetic evidence and found that seven proteins (B4GAT1, DNER, ENO3, HMOX2, OMG, RTBDN, TSPAN8) have effects on T2D, six proteins (CD34, FGFBP3, GALNT10, KHK, MENT, MXRA8) were affected by T2D, and five proteins (GSTA1, GSTA3, MEGF9, NCAN, SHBG) were bidirectionally associated with T2D. Three proteins (CD34, KCTD5, SCT) have MRT1 and CT1 evidence with iT2D (i.e., the most robust MR and colocalization evidence), however the directions of the MR and observational estimates with iT2D were inconsistent (**Supplementary Table 15**).

### China Kadoorie Biobank – EAS

In CKB-EAS, 315 and 416 proteins (483 of which were unique) were associated (p-value < 1.7 × 10^-5^; Bonferroni-corrected) with random blood glucose and pT2D, respectively. No protein was associated with iT2D at Bonferroni-corrected significance, while 10 proteins passed an FDR-corrected threshold < 0.05. In total, 484 proteins were associated with any of the three traits at this FDR-corrected threshold (**Supplementary Fig. 1** and **Supplementary Table 4**). Detailed observational estimates are in **Supplementary Table 12**.

As metabolic traits were not available in CKB-EAS (in participants with proteomics measured), we did not undertake bNMF clustering in CKB-EAS. Therefore, we focused on the 906 cluster defining proteins we identified in the UKB-EUR. Thirty-one proteins had too few pQTL in the CKB-EAS study and were excluded (**Supplementary Information**).

For the genetic analyses, we used previously published protein GWAS in CKB[33] (**Methods)**. In the first step, colocalization we tested whether the variants associated with protein levels were the same as those associated with diabetes or anthropometric traits. Ten (out of 878) proteins passed the “Relaxed Colocalization” (CT1 or CT2) and one (FST) passed the “Strict Colocalization” (CT1) with at least one of four diabetes traits (HbA1c, fasting glucose, fasting insulin, T2D). In the second step colocalization, only one protein (ENTR1) passed “Strict Colocalization” with at least one of the 17 cluster-specific metabolic/anthropometric traits (**Supplementary Table 13**). In summary, no protein was genetically aligned with both one diabetes trait and one metabolic/anthropometric trait passing the “Strict Colocation” criteria. Detailed colocalization results are provided in **Online Results**.

In MR analyses (i.e., nine (FST has no available *cis*-pQLT) robustly colocalised proteins with 23 traits (4 diabetes related traits and 19 cluster leading traits)), we found 17 MRT1 and 20 MRT2 protein – trait pairs. Incorporating the colocalization evidence, one pairs were in MRT1 and CT1 (ENTR1 – total protein); nine pairs were in MRT1/MRT2 and CT1/CT2. In reverse MR analyses, we found no MRT1 but 21 MRT2 trait – protein pairs. Supporting by colocalization evidence, three proteins (BOC, FGFBP3, PLA2G15) were in MRT1/MRT2 and CT1/CT2 affecting T2D (**Supplementary Table 17**), however the bidirectional effects were not supported by the MR Steiger directionality test (**Supplementary Table 16**). In sum, the genetic evidence suggested eight proteins affecting T2D (**Supplementary Table 15**). Statistical details of MR estimates are shown in **Online Results**.

We triangulated observational and genetic evidence, which suggested three proteins (NCR3LG1, RTBDN, ENTR1) affect T2D, with ENTR1 specific to CKB-EAS (**Supplementary Table 15**).

### Comparison of findings in UKB-EUR and CKB-EAS

In observational analyses, most (468) of the 484 proteins associated with either random glucose, pT2D or iT2D in CKB-EAS were supported in UKB-EUR (**Supplementary Fig. 1**) and the estimates from UKB-EUR and CKB-EAS were themselves correlated (ranging from 0.66 to 0.91) (**Supplementary Fig. 4**).

Sixteen proteins were only associated with T2D-relevant traits in the CKB-EAS population (FUCA1, GP2, LILRB5, TINAGL1, USP8, EDAR, VEGFD, CD99, RSPO3, CEACAM6, INSR, SERPINA6, SELENOP, TXNL1, NARS1, AMPD3).

Of the 54 proteins that shared causal genetic variants (i.e. colocalised with at least one diabetes trait) in UKB-EUR, six (BOC, FGFBP3, NCR3LG1, NELL1, RTBDN, TSPAN8) also had colocalization evidence with T2D in CKB-EAS. Moreover, four proteins (ACY1, ENTR1, FST, and PLA2G15) that colocalized with T2D in CKB-EAS, had weak colocalization evidence with T2D in UKB-EUR (**Supplementary Table 13**).

Genetic evidence (colocalization and MR) suggested that NCR3LG1, RTBDN, and TSPAN8 affected T2D in both populations (**Fig. 3** and **4**). Triangulating with observational evidence, the effects of RTBDN on T2D were consistent across populations. For TSPAN8, the observational and genetic estimates were directionally consistent, though the observational estimate with iT2D in CKB-EAS crossed the null. For NCR3LG1, apart from the observational estimate with iT2D in UKB-EUR crossed the null, others supported an inverse effect on T2D in both populations (**Table 2**).

**Fig. 3.**
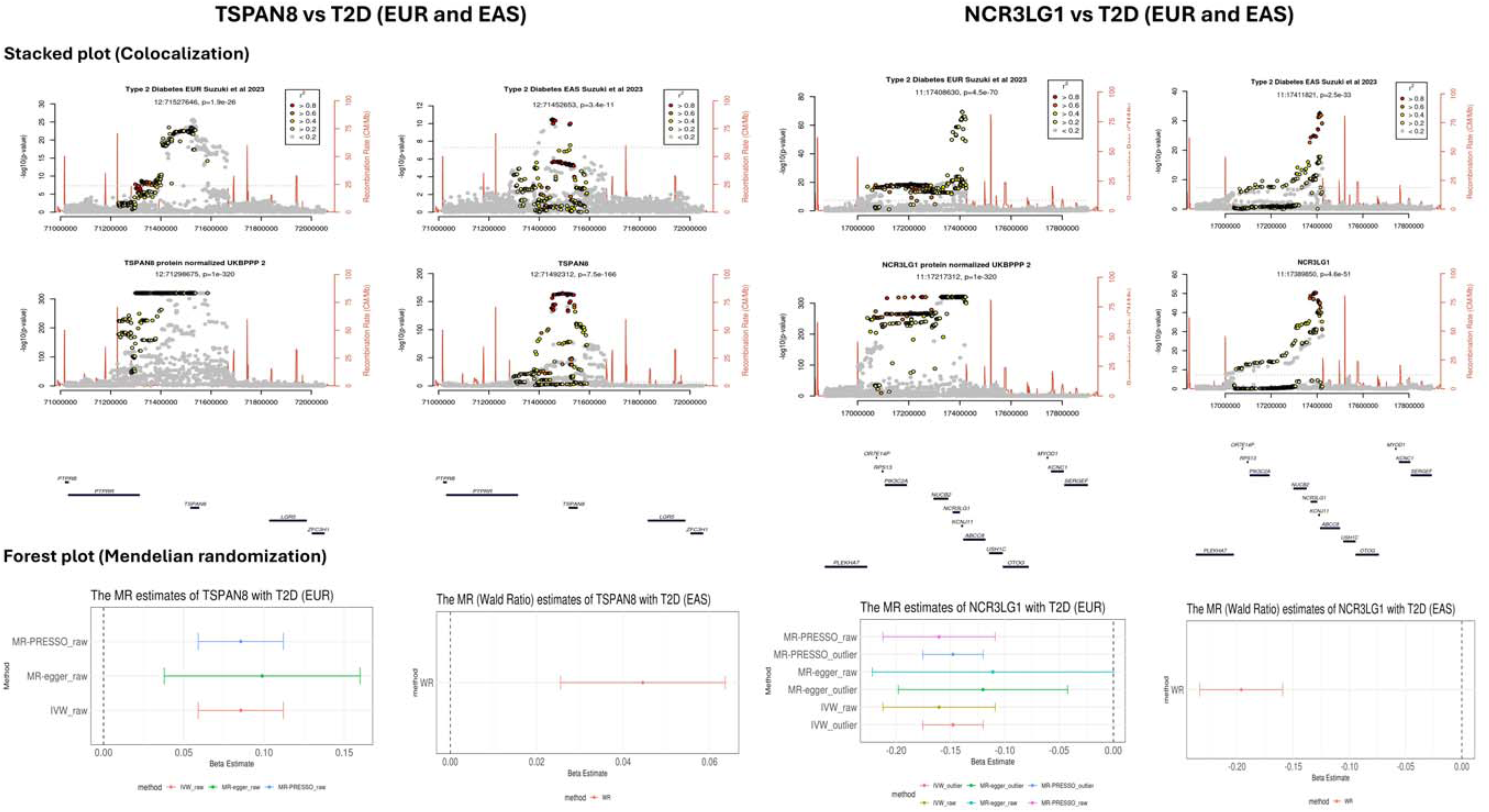
Colocalization (stacked plot) and Mendelian randomization (forest plot) estimates of TSPAN8 and NCR3LG1 with T2D risk in European and East Asian populations. TSPAN8: Tetraspanin-8, NCR3LG1: Natural cytotoxicity triggering receptor 3 ligand 1, T2D: type 2 diabetes, IVW: inverse variance weighted, MR-PRESSO: Mendelian randomization pleiotropy residual sum and outlier; “_raw”: the MR estimates without excluding outlier genetic variants; “_outlier”: the MR estimates excluding the outlier genetic variants, which were detected by the MR-PRESSO method.

**Fig. 4.**
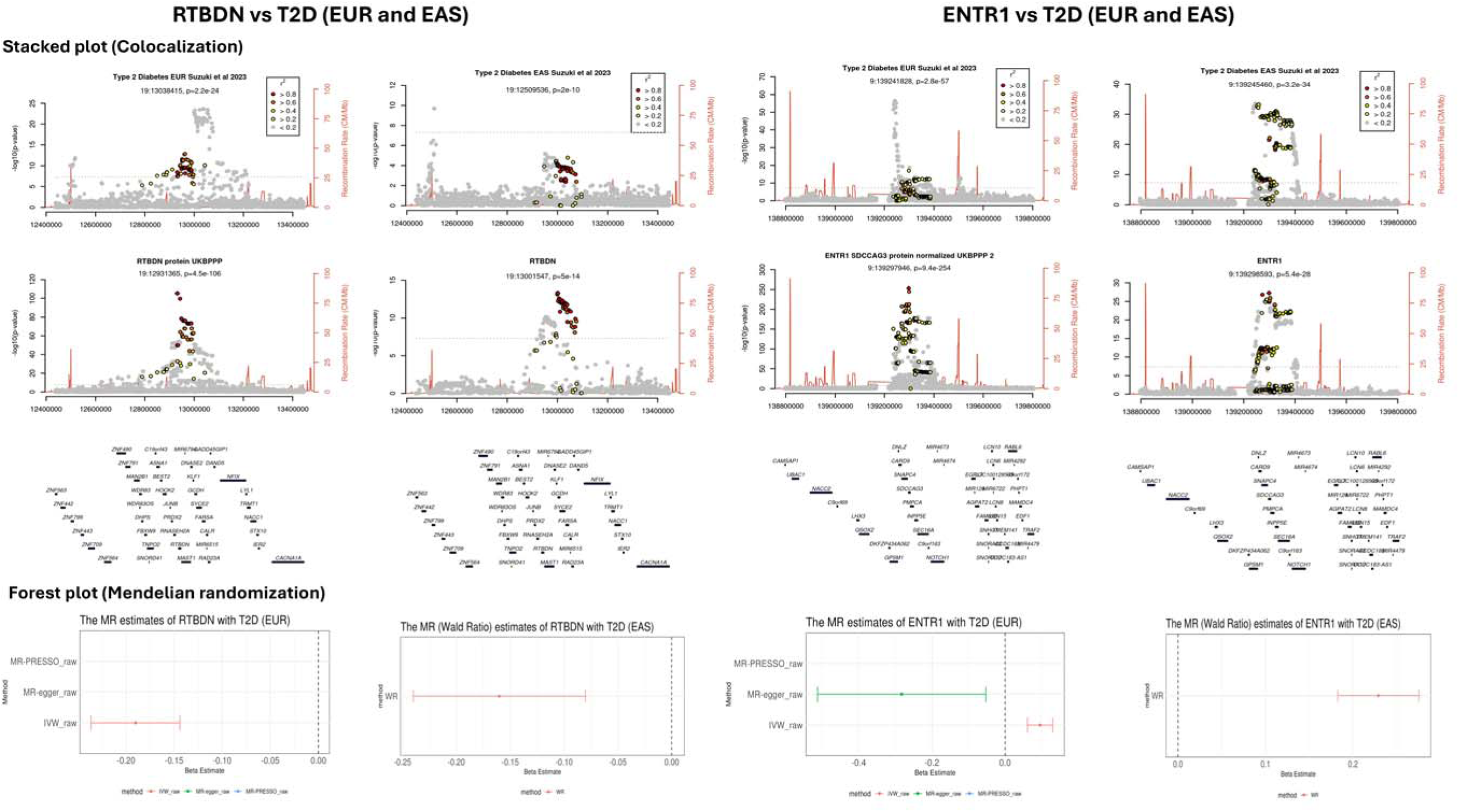
Colocalization (stacked plot) and Mendelian randomization (forest plot) estimates of RTBDN and ENTR1 with T2D risk in European and East Asian populations. RTBDN: Retbindin, ENTR1: Endosome-associated-trafficking regulator 1, T2D: type 2 diabetes, IVW: inverse variance weighted, MR-PRESSO: Mendelian randomization pleiotropy residual sum and outlier; “_raw”: the MR estimates without excluding outlier genetic variants; “_outlier”: the MR estimates excluding the outlier genetic variants, which were detected by the MR-PRESSO method.

**Table 2.**
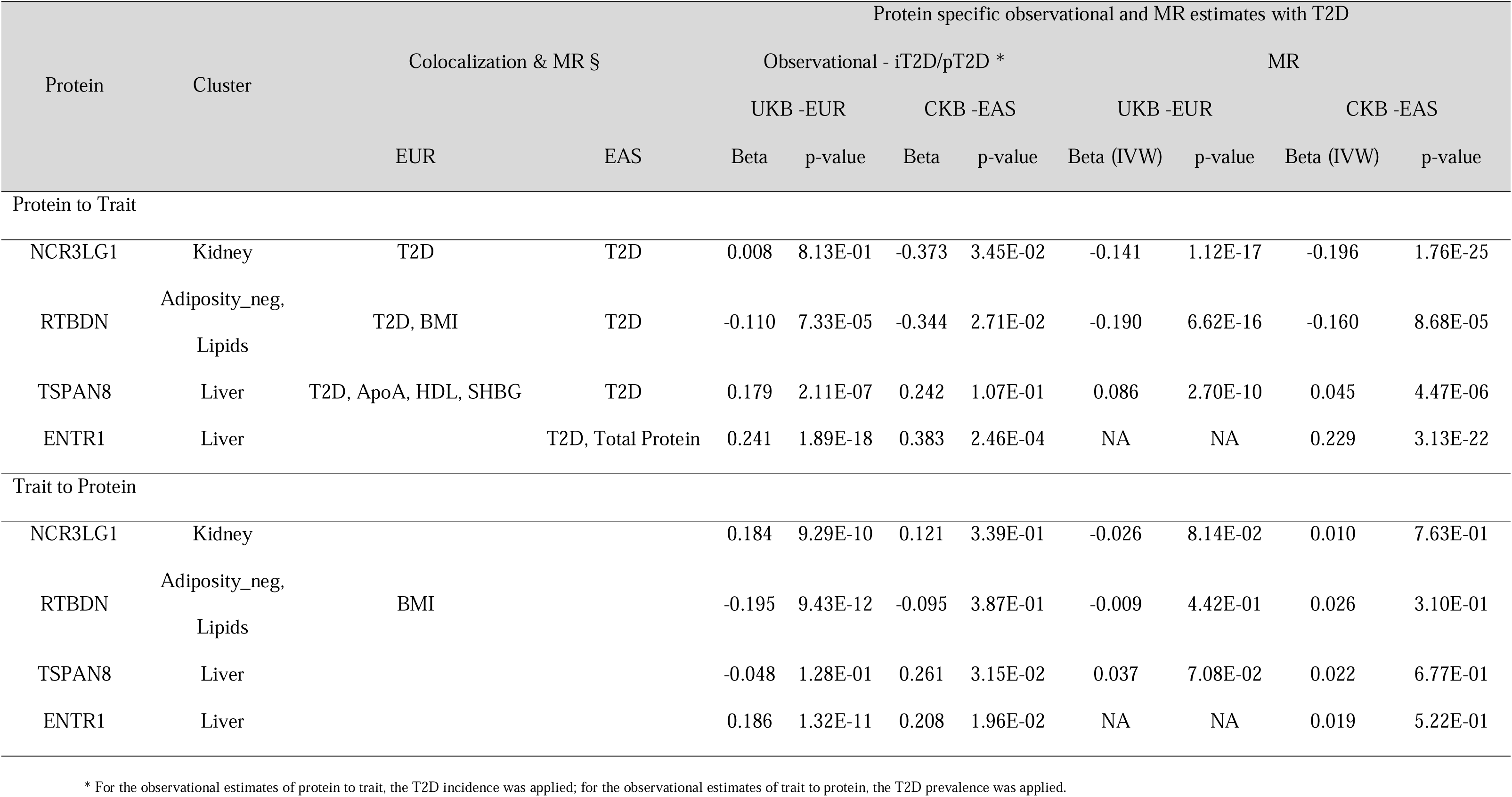
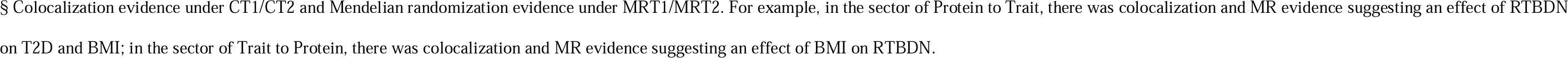
Genetic, observational, and Mendelian randomization evidence for four key T2D-associated proteins in European and East Asian populations.

## Discussion

To reveal heterogeneous aetiology (cluster specific) of T2D and identify potential drug targets, we identified and classified diabetes-associated proteins incorporating metabolic and anthropometric profiles and then leveraged genetic information to investigate the three-way causal relationships in two different ancestry populations. In UKB-EUR, we identified 1,793 diabetes-associated proteins and then classified into five clusters (Adiposity, Reduced-adiposity, Lipids, Liver, Kidney). Through triangulation of observational and genetic evidence, we found eight proteins affected T2D, six proteins were affected by T2D, and five proteins were bidirectionally associated with T2D. In CKB-EAS, four proteins affected T2D, including one (ENTR1) specific to CKB-EAS. The effects of RTBDN, TSPAN8, and NCR3LG1 on T2D were cross-ancestry consistent.

### Comparison with previous findings

Of the five clusters identified in our study, three (Adiposity, Lipids, Liver) were similar to those previously reported based on T2D clinical[4, 5] and genetic[6–9] profiles. However, we did not identify the beta-cell function and insulin clusters, likely due to the lack of glycaemic traits (e.g. HbA1c, fasting glucose) and beta-cell phenotypes (e.g. fasting insulin, HOMA, HOMA-B) in the clustering step. Moreover, the ∼3,000 measured proteins did not capture the full proteome, particularly those related to specific tissues and/or organs were not covered due to lack of detectability, specificity, or unknown clinical relevance. Future analyses should include tissue-specific proteomic data that may become more widely available. Combing proteins with metabolic and anthropometric profiles, we identified two new clusters (i.e., adiposity-reduced and kidney), which were not previously reported. The adiposity-reduced cluster (which assumes that higher levels of the protein associated with lower levels of adiposity traits and increased T2D risk) might provide insights into the T2D risk among individuals with lower BMI (e.g., EAS with young-onset T2D).[52, 53] This cluster could possibly reflect lipodystrophy features as recently discovered.[9] The genetic lipodystrophy cluster(s) reported by Smith et al[9] involving insulin resistance with traits related to lipid metabolism, body composition (increased visceral adipose tissue), and liver dysfunction, which was highly expressed in the EAS population. By accounting for lipodystrophy cluster-specific genetic score, the difference in T2D risk-equivalent BMI thresholds between the EAS and EUR ancestry groups decreased by approximately 4Dkg/m^2^ (i.e., EUR and EAS shared equivalent T2D risk at BMI of 30 and 24 kg/m^2^, respectively. Adjusting for the lipodystrophy genetic score the BMI threshold was increased to 28 kg/m^2^ in the EAS). The kidney cluster identified in our study was aligned with the well-established link between T2D and renal diseases, which was considered a common complication in diabetes patients being independent of glycaemic traits except for glucose on chronic kidney disease.[54]

The observational and genetic analyses in UKB-EUR identified 11 proteins (B4GAT1, DNER, ENO3, HMOX2, OMG, RTBDN, GSTA1, GSTA3, MEGF9, NCAN, SHBG) causally associated with T2D, which replicated the findings of an earlier proteome-wide MR study[55] except one protein (TSPAN8), which was not previously tested. For the five proteins with potential bidirectional effects, three (GSTA1, NCAN, SHBG) were previously shown with bidirectional MR evidence and two (GSTA3, MEGF9) were not previously tested.[55] Six (CD34, FGFBP3, GALNT10, KHK, MENT, MXRA8) proteins might be post-T2D markers (i.e., affected by T2D), though no directly evidence was found previously. Indirectly, three (MENT, MXRA8, CD34) of the six were not found to affect T2D.[55] Conversely, FGFBP3 and GALNT10 have been suggested to affect T2D.[55] CD34, KCTD5, and SCT had strongest genetic evidence of effects on T2D (fulfilling CT1 and MRT1), but there were directional inconsistencies between the observational and MR findings. Further studies are needed to clarify the effects of CD34, SCT, and KCTD5 on T2D (**Supplementary Table 15** and **Online Results**).

In CKB-EAS, we found three proteins (NCR3LG1, RTBDN, ENTR1) affecting T2D risk in both observational and genetic analyses. NCR3LG1 and RTBDN were consistent with UKB-EUR, but no colocalization evidence of ENTR1 with T2D. Yao et al found three proteins (ENTR1, LPL, PON3) having effects on T2D in EAS.[11] We replicated ENTR1 and found weak genetic evidence of LPL and PON3 with T2D in either UKB-EUR or CKB-EAS (**Online Results)**.

### Potential pathways linking candidate proteins and T2D

We found three proteins (RTBDN, NCR3LG1, and TSPAN8) associated with T2D across ancestries. RTBDN was found bidirectionally associated with BMI (**Supplementary Table 15**), indicating effects of RTBDN on T2D via adiposity change. Consistently, in the common metabolic diseases knowledge portal (CMDKP),[56] gene *RTBDN* was suggested as having a moderate Human Genetic Evidence (HuGE) score (i.e., a score quantifies genetic support for involvement of candidate gene in the diseases and traits available in CMDKP, based on several kinds of human genetic results)[57] with T2D and strong HuGE scores with BMI and haematological traits. However, apart from this, there was limited external evidence suggesting any effects of RTBDN on T2D via adiposity change but links between RTBDN with diabetic retinopathy.[55, 58–60] For NCR3LG1, its effect on T2D appeared to be mediated through adiposity,[61–63] which were consistent with our colocalization estimates, though our MR evidence was weak (e.g., colocalized with BMI (CT2); no MR evidence on NCR3LG1 affecting BMI). In CMDKP, strong HuGE scores of *NCR3LG1* with BMI, T2D, HbA1c, and glucose were found. The *NCR3LG1* gene region was shared by leptin levels and T2D,[64] suggesting a potential path of NCR3LG1 on T2D via adiposity change associated with leptin level. For TSPAN8, its gene region *TSPAN8/ LGR5* was found to be associated with T2D, though the *TSPAN8* knockout mouse models did not display apparent alterations in fasting insulin and glucose levels.[65] We found robust effects of TSPAN8 on SHBG (**Supplementary Table 15**), indicating that TSPAN8 and SHBG may affect T2D via a shared biological pathway. In line with our findings, very strong HuGE scores were found of *TSPAN8* associated with SHBG, T2D, as well as strong and moderate HuGE scores with HbA1c and fasting glucose, respectively. The three proteins with homogeneous effects on T2D in both UKB-EUR and CKB-EAS may reflect the shared genetic variants across ancestries supported by the high pairwise linkage disequilibrium statistics (D’) of the cis-pQTLs of the corresponding proteins (maximum D’ ranged from 0.96 (RTBDN) to 1 (NCR3LG1 and TSPAN8) (The D’ statistics were from LDlink (https://ldlink.nih.gov/?tab=home) (**Online Results**). ENTR1 had a CKB-EAS specific effect on T2D. The *ENTR1* gene has strong HuGE scores with T2D and HbA1c. Apart from this, it exclusively had a very strong HuGE score with fasting insulin, which was not shown in other candidate proteins. *ENTR1* was ubiquitously expressed including in Beta-cells, which could contribute to ancestry differences of T2D due to fasting insulin caused by beta-cell dysfunction given the prevalence of Beta cell dysfunction in EAS populations.[52, 53] In addition, strong HuGE scores of *ENTR1* with dysfunction traits (e.g., neutrophil, albumin, ALP) were found, which was in line with the EAS highly-expressed lipodystrophy genetic cluster (related to liver dysfunction).[9] These genetic evidence implied ENTR1 a key role in EAS T2D risk. These four novel proteins identified in our work could be considered as putative T2D drug targets. However, the elucidation of their underlying biological pathways will necessitate functional follow-up analyses.

Apart from the above-mentioned candidate proteins, our analyses also identified other novel candidate proteins revealing potential pathways supporting by genetic estimates (**Supplementary Table 15**). For example, DLK1, DNER, GRP, and HMOX2 were found genetically associated with adiposity traits and T2D. Consistently, animal and population evidence supported their associations with T2D mediated by body composition related to insulin responsiveness/resistance.[66–71] NCAN was found genetically associated with liver enzymes in this study. Its effect on liver damage was considered via liver inflammation and fibrosis in obesity-associated non-alcoholic fatty liver disease,[72] a potential cause of T2D.[73] Four proteins (GSTA1, GSTA3, NCAN, NFKB1) were found genetically associated with both liver and kidney traits, which might explain the links of T2D with liver and/or kidney diseases.[74–76] We found genetic association of ELN with cystatin C, supporting the observed ELN deficiency in chronic kidney disease.[77, 78]

### Strengths and limitations

This study has several strengths. We identified the proteins associated with different diabetes outcomes (i.e., HbA1c, glucose, pT2D, iT2D) in two large cohorts. The characteristics of diabetes associated proteins were captured in a multi-level profile via their associations with 43 metabolic/anthropometric traits. Our study is the first to apply a soft-clustering method (i.e., non-exclusive, a protein or a metabolic/anthropometric trait can be assigned into ≥1 clusters) to understand the role of circulating protein levels in diabetes. We systematically investigated the causal relationship of diabetes associated proteins with ≥30 diabetes- and cluster-specific traits using two statistical genetic approaches.[4–9] The tri-variate relationships between proteins, intermediate traits and T2D helped reveal different T2D aetiologies. To identify the most important candidate protein – trait pairs, we developed two-Tier systems for colocalization and MR respectively, leveraging all evidence rather than driven by one single estimate. In addition, we triangulated the observational and genetic evidence across ancestries.

The study also has limitations. First, due to the lack of metabolic trait in the CKB-EAS, we could not perform independent clustering in the CKB-EAS, which may reveal new or different features. There is no guarantee that in a different cohort (a different ancestry), the same proteins will contribute to the same functional clusters in the same way. Second, our findings relied entirely on circulating proteins, and referred to gene expression measured in bulk tissues, which may not reflect protein/transcript levels in the cell types/tissues most relevant to that gene in the context of T2D. Third, focusing on the top 10% proteins was intuitive for interpretation. Kim et al[7] picked up the leading traits in a statistical perspective (i.e., aggregated the weights of all the proteins plotting in descending order then fitted a line to the top 1% of the weights and another line to the bottom 80% of the weight), which was difficult to interpret practically (**Supplementary Information**). We compared the two methods and found both identified similar leading proteins (**Supplementary Fig. 5**) and traits (**Supplementary Table 18**). Fourth, the agreement between the bNMF clustering and the genetic estimates was not ideal, likely due to the different sources of bias in observational and genetic designs. Future work is needed to compare bNMF estimates using observational and genetic background. Fifth, there were limitations in the MR analysis. False positive estimates could be presented due to pleiotropy, though we applied pleiotropy robust methods (i.e., MR-Egger and MR-PRESSO) whenever possible. A low number of instrument would limit power to detect pleiotropy as well as a non-zero MR estimate with the use of MR-Egger.[79] There were limitations in the MR Tier system. The comparison of two protein - trait MR estimates within the same Tier was not straightforward when the two proteins were with different number of cis-pQTLs. For example, it would be over-conservative to a protein with larger number of cis-pQTLs given all the pleiotropy robust estimates were considered as well as accounting for Bonferroni corrected multiple-testing (e.g., 30 cis-pQTLs for NCR3L1 *vs* 1 cis-pQTL for SCT). Lastly, there were concerns of false positive in the reverse MR (trait to protein) given a comprehensive assessment of the credibility (e.g., inappropriate choice of genetic variants, insufficient interrogation of findings, violation of the relevance assumption taking disease as an exposure et al)[80, 81] were not practically available with massive amount of tests.

### Conclusion

This study identified 1,793 diabetes-associated proteins and five biological clusters related to T2D. Triangulating the observational and genetic evidence, we found a total of 20 T2D-associated causal proteins, including 19 in UKB-EUR and four in CKB-EAS (ENTR1 was CKB-EAS specific). Moreover, we revealed candidate proteins linked to T2D via adiposity, liver, and kidney traits, thus providing insights into different aetiology as well as links with diabetes comorbidities. Beyond their potential as therapeutic targets, these proteins represent promising biomarkers for T2D risk stratification, prognosis, and prediction of future disease progression and complications. Future work could prioritise testing the prognostic and predictive utility of these proteins in longitudinal proteomics datasets, assessing their ability to improve early detection, disease monitoring, and outcome prediction beyond established clinical risk factors. Further epidemiological and functional studies are required to replicate these findings, clarify underlying mechanisms, and translate them into actionable clinical applications.

## Declarations

### Ethics approval and consent to participate

All UK Biobank data are available to the research community via an application (reference number 65851). All participants provided informed consent.

For CKB, Prior to starting the project, central ethics approvals were obtained from Oxford University and the China National Center for Disease Control and Prevention (CDC). In addition, approvals were also obtained from institutional research boards at the local CDCs in the ten regions. All participants provided written informed consent

## Supporting information

Online Tables 1- 12, which contains large tables of the detailed estimates

## Data Availability

All UK Biobank data are available to the research community via an application, with full details of this process provided on the study website (https://www.ukbiobank.ac.uk/enable-your-research/apply-for-access).
The CKB is a global resource for the investigation of lifestyle, environmental, blood biochemical, and genetic factors as determinants of common diseases. The CKB study group is committed to making the cohort data available to the scientific community in China, the U.K., and worldwide to advance knowledge about the causes, prevention, and treatment of disease. For detailed information on what data are currently available to open access users and how to apply for it, please visit https://www.ckbiobank.org/data-access/data-access-procedures. A research proposal will be requested to ensure that any analysis is performed by bona fide researchers. Researchers who are interested in obtaining additional information or data that underlies this paper should contact. For any data that are not currently available for open access, researchers may need to develop formal collaboration with a study group. Custom code was used for all statistical analyses in this report.
R scripts for the key analyses are available on GitHub at: https://github.com/JamesLiu-OxfordNN/Proteomics_Diabetes_Clustering.git. Other statistical codes (e.g., colocalization and Mendelian randomization) were conducted with the pipeline and server in Novo Nordisk Research Centre of Oxford. Please contact the corresponding author via.

## Acknowledgements

Data on glycaemic traits have been contributed by MAGIC investigators and have been downloaded from www.magicinvestigators.org. We acknowledged Dr. Sile Hu who conducted the GWAS analysis of the UKB-PPP data in the Novo Nordisk Research Centre of Oxford. UKB data access reference number 65851 from Novo Nordisk.

## Funding

J.L. is supported by a Novo Nordisk Postdoctoral Fellowship run in partnership with the University of Oxford. CKB are supported by the grants from the National Natural Science Foundation of China (82192904, 82192903, 82192900,82388102, 82192901) and the Noncommunicable Chronic Diseases-National Science and Technology Major Project (2023ZD0510100) in China. In the UK, the UK Medical Research Council (MC_UU_00017/1,MC_UU_12026/2, MC_U137686851), Cancer Research UK (C16077/A29186; C500/A16896) and the British Heart Foundation (CH/1996001/9454), provide core funding to the Clinical Trial Service Unit and Epidemiological Studies Unit at Oxford University for the project. This research was funded in whole, or in part, by the Wellcome Trust (212946/Z/18/Z, 202922/Z/16/Z, 104085/Z/14/Z, and 088158/Z/09/Z). For the purpose of Open Access, the author has applied a CC-BY public copyright licence to any Author Accepted Manuscript version arising from this submission

## Authors’ contribution

J.L. conceived the idea for the study and carried out the data analysis and wrote the first draft of the manuscript. Z.C. and J.H. obtained funds, supervised, and jointly developed analysis plan with J.L. and provided critical input into manuscript writing, presentation and interpretation of the findings. L.C., R.N., N.R., M.T., A.P., L.B., and S.S. provided analytical supports (e.g., data access and statistical pipeline). All authors provided critical comments and revisions to the paper.

J.L., Z.C., and J.H. affirm that this manuscript is an honest, accurate, and transparent account of the study being conducted and reported, that no important aspects of the study have been omitted, and that any discrepancies from the study as planned (and, if relevant, registered) have been explained.

## Competing of interests

J.L. is supported by a Novo Nordisk Postdoctoral Fellowship run in partnership with the University of Oxford., J.H., L.C., R.N, N.R., M.T., S.S., W.G., and G.A. are full-time employees in Novo Nordisk. J.H., L.C., R.N, W.G., and G.A. are shareholders in Novo Nordisk. All other authors have no conflict of interest to declare.

## Availability of data and materials

The dataset(s) supporting the conclusions of this article is(are) available in the UK Biobank (UKB) and China Kadoorie Biobank (CKB).

All UK Biobank data are available to the research community via an application, with full details of this process provided on the study website (https://www.ukbiobank.ac.uk/enable-your-research/apply-for-access).

The CKB is a global resource for the investigation of lifestyle, environmental, blood biochemical, and genetic factors as determinants of common diseases. The CKB study group is committed to making the cohort data available to the scientific community in China, the U.K., and worldwide to advance knowledge about the causes, prevention, and treatment of disease. For detailed information on what data are currently available to open access users and how to apply for it, please visit https://www.ckbiobank.org/data-access/data-access-procedures. A research proposal will be requested to ensure that any analysis is performed by bona fide researchers. Researchers who are interested in obtaining additional information or data that underlies this paper should contact. For any data that are not currently available for open access, researchers may need to develop formal collaboration with a study group. Custom code was used for all statistical analyses in this report.

R scripts for the key analyses are available on GitHub at: https://github.com/JamesLiu-OxfordNN/Proteomics_Diabetes_Clustering.git. Other statistical codes (e.g., colocalization and Mendelian randomization) were conducted with the pipeline and server in Novo Nordisk Research Centre of Oxford. Please contact the corresponding author via

## List of Supplementary Materials

### Supplementary Tables

Supplementary Table 1 Summary information for publicly available genome-wide association studies included in the colocalization and Mendelian randomization analyses

Supplementary Table 2. Highly correlated metabolic/anthropometric traits (the square of phenotypic Pearson correlation coefficients >= 0.80) in UK Biobank

Supplementary Table 3. Highly correlated proteins (the square of phenotypic Pearson correlation coefficients >= 0.80) in UK Biobank

Supplementary Table 4 Number of proteins associated with glycaemic traits, diabetes prevalence, and diabetes incidence across observational designs and models in the UK Biobank (UKB-EUR) and China Kadoorie Biobank (CKB-EAS)

Supplementary Table 5 Leading features/traits of each cluster identified by bNMF across five diabetes outcomes, with and without adjustment for protein–protein and trait–trait correlations

Supplementary Table 6 Leading proteins of each cluster identified by bNMF across five diabetes outcomes, with and without adjustment for protein–protein and trait–trait correlations

Supplementary Table 7 Characteristics of genetic instruments (cis-pQLTs) for candidate proteins in the UK Biobank applied in the Mendelian randomization analysis

Supplementary Table 8 Characteristics of genetic instruments (cis-pQLTs) of candidate proteins in the China Kadoorie Biobank applied in the Mendelian randomization analysis

Supplementary Table 9 Colocalized robust proteins in the UK Biobank whose corresponding cis-pQTLs were not detected in the outcome-specific GWAS

Supplementary Table 10 Proxy genetic variants of cis-pQTLs for colocalized proteins in the China Kadoorie Biobank and missing outcome GWAS estimates due to variant unavailability

Supplementary Table 11 Criteria for Mendelian randomization estimation tiers

Supplementary Table 12 Observational estimates of all measured proteins with glycaemic traits, type 2 diabetes prevalence, and incidence in the UK Biobank (UKB-EUR) and China Kadoorie Biobank (CKB-EAS)

Supplementary Table 13 Fifty-eight diabetes-colocalized proteins and their colocalization evidence with cluster-specific traits (protein–trait pairs) in European and East Asian populations

Supplementary Table 14 Combined colocalization and bidirectional Mendelian randomization results (protein–trait pairs) for 54 diabetes-colocalized proteins, aligned with the corresponding colocalization and Mendelian randomization tier system in the European population

Supplementary Table 15 Observational and Mendelian randomization estimates of 45 diabetes-associated proteins (with robust genetic evidence) for type 2 diabetes in European and East Asian populations

Supplementary Table 16 Mendelian randomization Steiger directionality test for potential bidirectional protein–type 2 diabetes pairs in European and East Asian populations

Supplementary Table 17 Combined colocalization and bidirectional Mendelian randomization results (protein–trait pairs) for 10 diabetes-colocalized proteins, aligned with the corresponding colocalization and Mendelian randomization tier system in the East Asian population

Supplementary Table 18 Comparison of cluster-defining traits using the leading method versus the cut-off method

### Online Results/Tables

Online Table 1. Detailed colocalization estimates of diabetes-associated proteins with diabetes traits and cluster-leading metabolic/anthropometric traits in the European population

Online Table 2. Detailed colocalization estimates of diabetes-associated proteins with diabetes traits and cluster-leading metabolic/anthropometric traits in the East Asian population

Online Table 3. Detailed Mendelian randomization estimates of 53 diabetes-colocalized proteins with diabetes traits and cluster-leading metabolic/anthropometric traits (i.e., protein to trait) in the European population

Online Table 4. Detailed Mendelian randomization estimates of 9 diabetes-colocalized proteins with diabetes traits and cluster-leading metabolic/anthropometric traits (i.e., protein to trait) in the East Asian population

Online Table 5. Detailed Mendelian randomization estimates of diabetes traits and cluster-leading metabolic/anthropometric traits with 54 diabetes-colocalized proteins (i.e., trait to protein) in the European population

Online Table 6. Detailed Mendelian randomization estimates of diabetes traits and cluster-leading metabolic/anthropometric traits with 10 diabetes-colocalized proteins (i.e., trait to protein) in the East Asian population

Online Table 7. Pairwise linkage disequilibrium statistics (R^2^ and D’) of cis-pQTLs for RTBDN in UK Biobank and China Kadoorie Biobank (1000 Genomes EUR reference panel)

Online Table 8. Pairwise linkage disequilibrium statistics (R^2^ and D’) of the cis-pQTLs for RTBDN in UK Biobank and China Kadoorie Biobank (1000 Genome, EAS as the reference panel)

Online Table 9. Pairwise linkage disequilibrium statistics (R^2^ and D’) of the cis-pQTLs for NCR3LG1 in UK Biobank and China Kadoorie Biobank (1000 Genomes EUR reference panel)

Online Table 10. Pairwise linkage disequilibrium statistics (R^2^ and D’) of the cis-pQTLs for NCR3LG1 in UK Biobank and China Kadoorie Biobank (1000 Genome, EAS as the reference panel)

Online Table 11. Pairwise linkage disequilibrium statistics (R^2^ and D’) of the cis-pQTLs for TSPAN8 in UK Biobank and China Kadoorie Biobank (1000 Genomes EUR reference panel)

Online Table 12. Pairwise linkage disequilibrium statistics (R^2^ and D’) of the cis-pQTLs for TSPAN8 in UK Biobank and China Kadoorie Biobank (1000 Genome, EAS as the reference panel)

### Supplementary Figures

Supplementary Fig. 1. Number of overlapping leading proteins across all bNMF models (e.g., HbA1c vs glucose outcomes; with and without protein–protein and trait–trait correlations)

Supplementary Fig. 2. Number of overlapped proteins associated with glycaemic traits and diabetes in (a) UK Biobank (UKB-EUR), (b) China Kadoorie Biobank (CKB-EAS) and (c) the proteins across the two cohorts

Supplementary Fig. 3. Colocalization (stacked plot) and Mendelian randomization (forest plot) estimates of CD34, KCTD5, and SCT with T2D risk in European population

Supplementary Fig. 4. Correlations of the observational estimates (protein–diabetes trait associations) between UK Biobank (UKB-EUR) and China Kadoorie Biobank (CKB-EAS)

Supplementary Fig. 5. Number of overlapped cluster-leading proteins using the leading method and the cut-off method

## Notes

### Competing Interest Statement

The authors have declared no competing interest.

### Author Declarations

All UK Biobank data are available to the research community via an application, with full details of this process provided on the study website (https://www.ukbiobank.ac.uk/enable-your-research/apply-for-access). The CKB is a global resource for the investigation of lifestyle, environmental, blood biochemical, and genetic factors as determinants of common diseases. The CKB study group is committed to making the cohort data available to the scientific community in China, the U.K., and worldwide to advance knowledge about the causes, prevention, and treatment of disease. For detailed information on what data are currently available to open access users and how to apply for it, please visit https://www.ckbiobank.org/data-access/data-access-procedures. A research proposal will be requested to ensure that any analysis is performed by bona fide researchers. Researchers who are interested in obtaining additional information or data that underlies this paper should contact. For any data that are not currently available for open access, researchers may need to develop formal collaboration with a study group. Custom code was used for all statistical analyses in this report.

### Summary of Updates

A Graphic abstract and the clarification for Supplementary Figure 4 have been added. Minor typographical (e.g., "HOMX2" is revised as "HMOX2") and formatting errors have been corrected, and the text and table names have been edited to improve clarity. No substantive changes were made to the analyses, and the findings and conclusions remain unchanged.

## References

1. Ong KL, Stafford LK, McLaughlin SA, Boyko EJ, Vollset SE, Smith AE, Dalton BE, Duprey J, Cruz JA, Hagins H et al: Global, regional, and national burden of diabetes from 1990 to 2021, with projections of prevalence to 2050: a systematic analysis for the Global Burden of Disease Study 2021. The Lancet 2023, 402(10397):203–234.

2. Cannon A, Handelsman Y, Heile M, Shannon M: Burden of Illness in Type 2 Diabetes Mellitus. J Manag Care Spec Pharm 2018, 24(9-a Suppl):S5–s13.

3. Committee ADAPP: 9. Pharmacologic Approaches to Glycemic Treatment: Standards of Care in Diabetes—2024. Diabetes Care 2023, 47(Supplement_1):S158–S178.

4. Christensen DH, Nicolaisen SK, Ahlqvist E, Stidsen JV, Nielsen JS, Hojlund K, Olsen MH, García-Calzón S, Ling C, Rungby J et al: Type 2 diabetes classification: a data-driven cluster study of the Danish Centre for Strategic Research in Type 2 Diabetes (DD2) cohort. BMJ Open Diabetes Res Care 2022, 10(2).

5. Wesolowska-Andersen A, Brorsson CA, Bizzotto R, Mari A, Tura A, Koivula R, Mahajan A, Vinuela A, Tajes JF, Sharma S et al: Four groups of type 2 diabetes contribute to the etiological and clinical heterogeneity in newly diagnosed individuals: An IMI DIRECT study. Cell Reports Medicine 2022, 3(1):100477.

6. Udler MS, Kim J, von Grotthuss M, Bonàs-Guarch S, Cole JB, Chiou J, Boehnke M, Laakso M, Atzmon G, Glaser B et al: Type 2 diabetes genetic loci informed by multi-trait associations point to disease mechanisms and subtypes: A soft clustering analysis. PLoS Med 2018, 15(9):e1002654.

7. Kim H, Westerman KE, Smith K, Chiou J, Cole JB, Majarian T, von Grotthuss M, Kwak SH, Kim J, Mercader JM et al: High-throughput genetic clustering of type 2 diabetes loci reveals heterogeneous mechanistic pathways of metabolic disease. Diabetologia 2023, 66(3):495–507.

8. Suzuki K, Hatzikotoulas K, Southam L, Taylor HJ, Yin X, Lorenz KM, Mandla R, Huerta-Chagoya A, Melloni GEM, Kanoni S et al: Genetic drivers of heterogeneity in type 2 diabetes pathophysiology. Nature 2024, 627(8003):347–357.

9. Smith K, Deutsch AJ, McGrail C, Kim H, Hsu S, Huerta-Chagoya A, Mandla R, Schroeder PH, Westerman KE, Szczerbinski L et al: Multi-ancestry polygenic mechanisms of type 2 diabetes. Nat Med 2024, 30(4):1065–1074.

10. Sun BB, Chiou J, Traylor M, Benner C, Hsu Y-H, Richardson TG, Surendran P, Mahajan A, Robins C, Vasquez-Grinnell SG et al: Plasma proteomic associations with genetics and health in the UK Biobank. Nature 2023, 622(7982):329–338.

11. Yao P, Iona A, Pozarickij A, Said S, Wright N, Lin K, Millwood I, Fry H, Kartsonaki C, Mazidi M et al: Proteomic Analyses in Diverse Populations Improved Risk Prediction and Identified New Drug Targets for Type 2 Diabetes. Diabetes Care 2024, 47(6):1012–1019.

12. Yao H, Zhang A, Li D, Wu Y, Wang C-Z, Wan J-Y, Yuan C-S: Comparative effectiveness of GLP-1 receptor agonists on glycaemic control, body weight, and lipid profile for type 2 diabetes: systematic review and network meta-analysis. BMJ 2024, 384:e076410.

13. van Baar MJB, van Ruiten CC, Muskiet MHA, van Bloemendaal L, IJzerman RG, van Raalte DH: SGLT2 Inhibitors in Combination Therapy: From Mechanisms to Clinical Considerations in Type 2 Diabetes Management. Diabetes Care 2018, 41(8):1543–1556.

14. Zheng J, Haberland V, Baird D, Walker V, Haycock PC, Hurle MR, Gutteridge A, Erola P, Liu Y, Luo S et al: Phenome-wide Mendelian randomization mapping the influence of the plasma proteome on complex diseases. Nature Genetics 2020, 52(10):1122–1131.

15. Zuber V, Grinberg NF, Gill D, Manipur I, Slob EAW, Patel A, Wallace C, Burgess S: Combining evidence from Mendelian randomization and colocalization: Review and comparison of approaches. Am J Hum Genet 2022, 109(5):767–782.

16. Burgess S, Mason AM, Grant AJ, Slob EAW, Gkatzionis A, Zuber V, Patel A, Tian H, Liu C, Haynes WG et al: Using genetic association data to guide drug discovery and development: Review of methods and applications. The American Journal of Human Genetics 2023, 110(2):195–214.

17. Sudlow C, Gallacher J, Allen N, Beral V, Burton P, Danesh J, Downey P, Elliott P, Green J, Landray M et al: UK biobank: an open access resource for identifying the causes of a wide range of complex diseases of middle and old age. PLoS Med 2015, 12(3):e1001779.

18. Fry A, Littlejohns TJ, Sudlow C, Doherty N, Adamska L, Sprosen T, Collins R, Allen NE: Comparison of Sociodemographic and Health-Related Characteristics of UK Biobank Participants With Those of the General Population. Am J Epidemiol 2017, 186(9):1026–1034.

19. Carter AR, Sanderson E, Hammerton G, Richmond RC, Davey Smith G, Heron J, Taylor AE, Davies NM, Howe LD: Mendelian randomisation for mediation analysis: current methods and challenges for implementation. Eur J Epidemiol 2021, 36(5):465–478.

20. Townsend P, Phillimore P, Beattie A: Health and deprivation: inequality and the North: Routledge; 1988.

21. UK Biobank Biomarker data [https://www.ukbiobank.ac.uk/enable-your-research/about-our-data/biomarker-data]

22. Bycroft C, Freeman C, Petkova D, Band G, Elliott LT, Sharp K, Motyer A, Vukcevic D, Delaneau O, O’Connell J et al: The UK Biobank resource with deep phenotyping and genomic data. Nature 2018, 562(7726):203–209.

23. UK Biobank Showcase [https://biobank.ndph.ox.ac.uk/showcase/]

24. Astle WJ, Elding H, Jiang T, Allen D, Ruklisa D, Mann AL, Mead D, Bouman H, Riveros-Mckay F, Kostadima MA et al: The Allelic Landscape of Human Blood Cell Trait Variation and Links to Common Complex Disease. Cell 2016, 167(5):1415–1429.e1419.

25. Biobank U: UK Biobank Olink proteomics data. In.; 2023.

26. Allen NE, Arnold M, Parish S, Hill M, Sheard S, Callen H, Fry D, Moffat S, Gordon M, Welsh S et al: Approaches to minimising the epidemiological impact of sources of systematic and random variation that may affect biochemistry assay data in UK Biobank. Wellcome Open Res 2020, 5:222.

27. Biobank U: UK Biobank First Occurrence of Health Outcomes Defined by 3-character ICD10 code. In.; 2019.

28. Chen Z, Chen J, Collins R, Guo Y, Peto R, Wu F, Li L: China Kadoorie Biobank of 0.5 million people: survey methods, baseline characteristics and long-term follow-up. Int J Epidemiol 2011, 40(6):1652–1666.

29. Walters RG, Millwood IY, Lin K, Schmidt Valle D, McDonnell P, Hacker A, Avery D, Edris A, Fry H, Cai N et al: Genotyping and population characteristics of the China Kadoorie Biobank. Cell Genomics 2023, 3(8):100361.

30. CKB data showcase contents [https://www.ckbiobank.org/datashowcase/18.01/showcase_contents.html]

31. Mbatchou J, Barnard L, Backman J, Marcketta A, Kosmicki JA, Ziyatdinov A, Benner C, O’Dushlaine C, Barber M, Boutkov B et al: Computationally efficient whole-genome regression for quantitative and binary traits. Nat Genet 2021, 53(7):1097–1103.

32. Purcell S, Neale B, Todd-Brown K, Thomas L, Ferreira MA, Bender D, Maller J, Sklar P, De Bakker PI, Daly MJ: PLINK: a tool set for whole-genome association and population-based linkage analyses. The American journal of human genetics 2007, 81(3):559–575.

33. Pozarickij A, Said S, Lin K, Morris S, Edris A, Iona A, Kartsonaki C, Wright N, Yao P, Fry H et al: Ancestry diversity in the genetic determinants of the human plasma proteome enhances identification of potential drug targets. medRxiv 2025:2023.2011.2013.23298365.

34. Suzuki K, Hatzikotoulas K, Southam L, Taylor HJ, Yin X, Lorenz KM, Mandla R, Huerta-Chagoya A, Melloni GEM, Kanoni S et al: Genetic drivers of heterogeneity in type 2 diabetes pathophysiology. Nature 2024, 627(8003):347–357.

35. Chen J, Spracklen CN, Marenne G, Varshney A, Corbin LJ, Luan J, Willems SM, Wu Y, Zhang X, Horikoshi M et al: The trans-ancestral genomic architecture of glycemic traits. Nat Genet 2021, 53(6):840–860.

36. Lagou V, Jiang L, Ulrich A, Zudina L, González KSG, Balkhiyarova Z, Faggian A, Maina JG, Chen S, Todorov PV et al: GWAS of random glucose in 476,326 individuals provide insights into diabetes pathophysiology, complications and treatment stratification. Nat Genet 2023, 55(9):1448–1461.

37. Broadaway KA, Yin X, Williamson A, Parsons VA, Wilson EP, Moxley AH, Vadlamudi S, Varshney A, Jackson AU, Ahuja V et al: Loci for insulin processing and secretion provide insight into type 2 diabetes risk. Am J Hum Genet 2023, 110(2):284–299.

38. Williamson A, Norris DM, Yin X, Broadaway KA, Moxley AH, Vadlamudi S, Wilson EP, Jackson AU, Ahuja V, Andersen MK et al: Genome-wide association study and functional characterization identifies candidate genes for insulin-stimulated glucose uptake. Nat Genet 2023, 55(6):973–983.

39. Chen CY, Chen TT, Feng YA, Yu M, Lin SC, Longchamps RJ, Wang SH, Hsu YH, Yang HI, Kuo PH et al: Analysis across Taiwan Biobank, Biobank Japan, and UK Biobank identifies hundreds of novel loci for 36 quantitative traits. Cell Genom 2023, 3(12):100436.

40. Pulit SL, Stoneman C, Morris AP, Wood AR, Glastonbury CA, Tyrrell J, Yengo L, Ferreira T, Marouli E, Ji Y et al: Meta-analysis of genome-wide association studies for body fat distribution in 694 649 individuals of European ancestry. Hum Mol Genet 2019, 28(1):166–174.

41. Pan-ancestry genetic analysis of the UK Biobank [https://pan.ukbb.broadinstitute.org/]

42. Sakaue S, Kanai M, Tanigawa Y, Karjalainen J, Kurki M, Koshiba S, Narita A, Konuma T, Yamamoto K, Akiyama M et al: A cross-population atlas of genetic associations for 220 human phenotypes. Nature Genetics 2021, 53(10):1415–1424.

43. Munafò MR, Tilling K, Taylor AE, Evans DM, Davey Smith G: Collider scope: when selection bias can substantially influence observed associations. International Journal of Epidemiology 2017, 47(1):226–235.

44. Foley CN, Staley JR, Breen PG, Sun BB, Kirk PDW, Burgess S, Howson JMM: A fast and efficient colocalization algorithm for identifying shared genetic risk factors across multiple traits. Nat Commun 2021, 12(1):764.

45. Wallace C: Eliciting priors and relaxing the single causal variant assumption in colocalisation analyses. PLOS Genetics 2020, 16(4):e1008720.

46. Burgess S, Thompson SG: Avoiding bias from weak instruments in Mendelian randomization studies. Int J Epidemiol 2011, 40(3):755–764.

47. Burgess SB, Jack: Integrating summarized data from multiple genetic variants in Mendelian randomization: bias and coverage properties of inverse-variance weighted methods. 2015.

48. Bowden J, Davey Smith G, Burgess S: Mendelian randomization with invalid instruments: effect estimation and bias detection through Egger regression. Int J Epidemiol 2015, 44(2):512–525.

49. Verbanck M, Chen C-Y, Neale B, Do R: Detection of widespread horizontal pleiotropy in causal relationships inferred from Mendelian randomization between complex traits and diseases. Nature Genetics 2018, 50(5):693–698.

50. Bowden J, Davey Smith G, Haycock PC, Burgess S: Consistent Estimation in Mendelian Randomization with Some Invalid Instruments Using a Weighted Median Estimator. Genet Epidemiol 2016, 40(4):304–314.

51. Hemani G, Tilling K, Davey Smith G: Orienting the causal relationship between imprecisely measured traits using GWAS summary data. PLOS Genetics 2017, 13(11):e1007081.

52. Fan Y, Chow E, Lim CKP, Hou Y, Tsoi STF, Fan B, Lau ESH, Kong APS, Ma RCW, Wu H et al: Comparison of β-Cell Function and Insulin Sensitivity Between Normal-Weight and Obese Chinese With Young-Onset Type 2 Diabetes. Diabetes 2024, 73(6):953–963.

53. Ma RC, Chan JC: Type 2 diabetes in East Asians: similarities and differences with populations in Europe and the United States. Ann N Y Acad Sci 2013, 1281(1):64–91.

54. Zheng J, Zhang Y, Rasheed H, Walker V, Sugawara Y, Li J, Leng Y, Elsworth B, Wootton RE, Fang S et al: Trans-ethnic Mendelian-randomization study reveals causal relationships between cardiometabolic factors and chronic kidney disease. International Journal of Epidemiology 2021, 50(6):1995–2010.

55. Yuan S, Xu F, Li X, Chen J, Zheng J, Mantzoros CS, Larsson SC: Plasma proteins and onset of type 2 diabetes and diabetic complications: Proteome-wide Mendelian randomization and colocalization analyses. Cell Rep Med 2023, 4(9):101174.

56. Common Metabolic Diseases Knowledge Portal [https://hugeamp.org/]

57. Dornbos P, Singh P, Jang DK, Mahajan A, Biddinger SB, Rotter JI, McCarthy MI, Flannick J: Evaluating human genetic support for hypothesized metabolic disease genes. Cell Metab 2022, 34(5):661–666.

58. Zhao X, Tebbe L, Naash MI, Al-Ubaidi MR: The Neuroprotective Role of Retbindin, a Metabolic Regulator in the Neural Retina. Front Pharmacol 2022, 13:919667.

59. Bhattacharya S, Kalra S, Dutta D, Khandelwal D, Singla R: The Interplay Between Pituitary Health and Diabetes Mellitus - The Need for ‘Hypophyseo-Vigilance’. Eur Endocrinol 2020, 16(1):25–31.

60. Cai L, Sun Y, Liu Y, Chen W, He L, Wei DQ: Evidence that the pituitary gland connects type 2 diabetes mellitus and schizophrenia based on large-scale trans-ethnic genetic analyses. J Transl Med 2022, 20(1):501.

61. Loesch DP, Garg M, Matelska D, Vitsios D, Jiang X, Ritchie SC, Sun BB, Runz H, Whelan CD, Holman RR et al: Identification of plasma proteomic markers underlying polygenic risk of type 2 diabetes and related comorbidities. medRxiv 2024:2024.2003.2015.24304200.

62. Coral DE, Fernandez-Tajes J, Tsereteli N, Pomares-Millan H, Fitipaldi H, Mutie PM, Atabaki-Pasdar N, Kalamajski S, Poveda A, Miller-Fleming TW et al: A phenome-wide comparative analysis of genetic discordance between obesity and type 2 diabetes. Nature Metabolism 2023, 5(2):237–247.

63. Maina JG, Balkhiyarova Z, Nouwen A, Pupko I, Ulrich A, Boissel M, Bonnefond A, Froguel P, Khamis A, Prokopenko I et al: Bidirectional Mendelian Randomization and Multiphenotype GWAS Show Causality and Shared Pathophysiology Between Depression and Type 2 Diabetes. Diabetes Care 2023, 46(9):1707–1714.

64. Wang X, Jia J, Huang T: Shared genetic architecture and casual relationship between leptin levels and type 2 diabetes: large-scale cross-trait meta-analysis and Mendelian randomization analysis. BMJ Open Diabetes Research & Care 2020, 8(1):e001140.

65. Yang J, Zhang Z, Lam JSW, Fan H, Fu NY: Molecular Regulation and Oncogenic Functions of TSPAN8. Cells 2024, 13(2).

66. Jensen CH, Kosmina R, Rydén M, Baun C, Hvidsten S, Andersen MS, Christensen LL, Gastaldelli A, Marraccini P, Arner P et al: The imprinted gene Delta like non-canonical notch ligand 1 (Dlk1) associates with obesity and triggers insulin resistance through inhibition of skeletal muscle glucose uptake. EBioMedicine 2019, 46:368–380.

67. Park J-R, Jung J-W, Seo M-S, Kang S-K, Lee Y-S, Kang K-S: DNER modulates adipogenesis of human adipose tissue-derived mesenchymal stem cells via regulation of cell proliferation. Cell Proliferation 2010, 43(1):19–28.

68. Ruiz-Otero N, Kuruvilla R: Role of Delta/Notch-like EGF-related receptor in blood glucose homeostasis. Front Endocrinol (Lausanne) 2023, 14:1161085.

69. Kim MK, Park HJ, Kim Y, Bae SK, Kim HJ, Bae MK: Involvement of Gastrin-Releasing Peptide Receptor in the Regulation of Adipocyte Differentiation in 3T3-L1 Cells. Int J Mol Sci 2018, 19(12).

70. Pendharkar SA, Drury M, Walia M, Korc M, Petrov MS: Gastrin-Releasing Peptide and Glucose Metabolism Following Pancreatitis. Gastroenterology Res 2017, 10(4):224–234.

71. Yao H, Peterson AL, Li J, Xu H, Dennery PA: Heme Oxygenase 1 and 2 Differentially Regulate Glucose Metabolism and Adipose Tissue Mitochondrial Respiration: Implications for Metabolic Dysregulation. Int J Mol Sci 2020, 21(19).

72. Gorden A, Yang R, Yerges-Armstrong LM, Ryan KA, Speliotes E, Borecki IB, Harris TB, Chu X, Wood GC, Still CD et al: Genetic variation at NCAN locus is associated with inflammation and fibrosis in non-alcoholic fatty liver disease in morbid obesity. Hum Hered 2013, 75(1):34–43.

73. Au Yeung SL, Borges MC, Wong THT, Lawlor DA, Schooling CM: Evaluating the role of non-alcoholic fatty liver disease in cardiovascular diseases and type 2 diabetes: a Mendelian randomization study in Europeans and East Asians. International Journal of Epidemiology 2022, 52(3):921–931.

74. Perdomo CM, Garcia-Fernandez N, Escalada J: Diabetic Kidney Disease, Cardiovascular Disease and Non-Alcoholic Fatty Liver Disease: A New Triumvirate? J Clin Med 2021, 10(9).

75. Orlić L, Mikolasevic I, Bagic Z, Racki S, Stimac D, Milic S: Chronic Kidney Disease and Nonalcoholic Fatty Liver Disease—Is There a Link? Gastroenterology Research and Practice 2014, 2014(1):847539.

76. Junghare MY, Ibrahim HN: Chapter 45 - Chronic Kidney Disease and Liver Disease. In: Chronic Renal Disease. Edited by Kimmel PL, Rosenberg ME. San Diego: Academic Press; 2015: 544–559.

77. Blaise S, Romier B, Kawecki C, Ghirardi M, Rabenoelina F, Baud S, Duca L, Maurice P, Heinz A, Schmelzer CEH et al: Elastin-Derived Peptides Are New Regulators of Insulin Resistance Development in Mice. Diabetes 2013, 62(11):3807–3816.

78. Wagenseil JE: Does elastin deficiency cause chronic kidney disease? Kidney International 2017, 92(5):1036–1038.

79. Burgess S, Thompson SG: Interpreting findings from Mendelian randomization using the MR-Egger method. Eur J Epidemiol 2017, 32(5):377–389.

80. Burgess S, Woolf B, Mason AM, Ala-Korpela M, Gill D: Addressing the credibility crisis in Mendelian randomization. BMC Medicine 2024, 22(1):374.

81. Chen S, Liang Y, Mo JMY, Li QHY, He B, Luo S, Burgess S, Au Yeung SL: Challenges in interpreting Mendelian randomization studies with a disease as the exposure: Using COVID-19 liability studies as an exemplar. European Journal of Human Genetics 2025.

