## Supplementary Figures 1 - 5 for "Identifying molecular pathways of type 2 diabetes using proteomics, metabolic, and anthropometric profiles in UK and Chinese adults"

**Supplementary Fig. 1. Number of overlapping leading proteins across all bNMF models (e.g., HbA1c vs glucose outcomes; with and without protein–protein and trait–trait correlations)**

bNMF clustering with the diabetes traits specific proteins (p-value < 1.7 × 10^-5; e.g., 1,466 proteins for HbA1c, 708 proteins for glucose; 787 proteins for diabetes prevalence; 815 proteins for T2D incidence; 1793 proteins for all diabetes outcomes)

We accounted for the trait – trait and protein – protein correlations, where the highly correlated (i.e., the square of Pearson correlation coefficients >= 0.80) traits (n = 8) and proteins (n = 182) were excluded

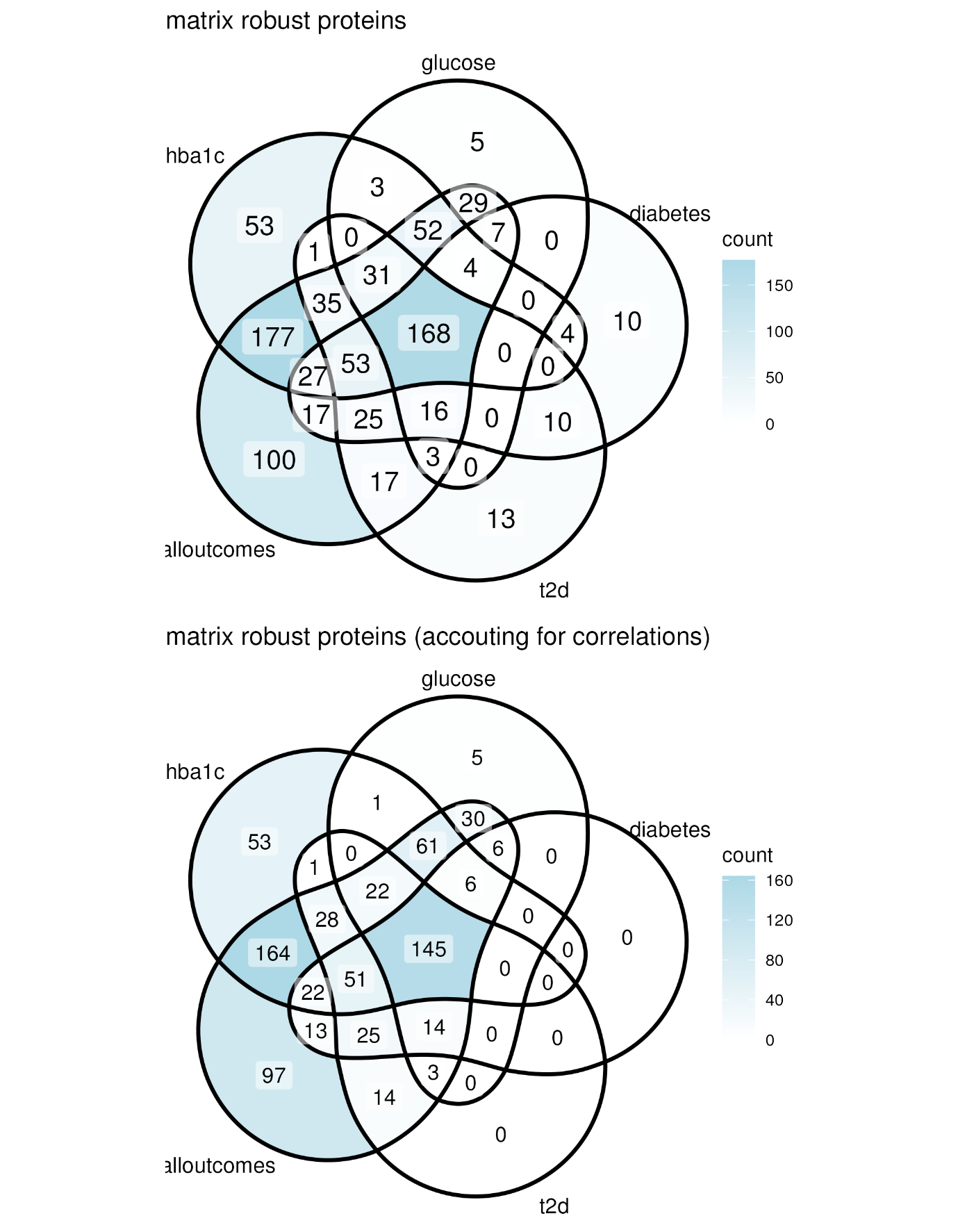

**Supplementary Fig. 2. Number of overlapped proteins associated with glycaemic traits and diabetes in (a) UK Biobank (UKB-EUR), (b) China Kadoorie Biobank (CKB-EAS) and (c) the proteins across the two cohorts**

In UKB-EUR, 1466, 708, 787, 815 proteins were associated (p-value < 1.7 × 10^^-5^) with HbA1c, random glucose, self-reported prevalent type 2 diabetes (pT2D), and T2D incidence (iT2D) respectively in both two models; among which, 315 proteins were associated with all four outcomes and 1,793 proteins were associated with at least one trait.

In CKB-EAS, 315 and 416 proteins were associated (p-value < 1.7 × 10^^-5^) with random glucose and self-reported pT2D in both two models. No protein was associated with Ti2D under a Bonferroni-corrected threshold (p-value < 1.7 × 10^^-5^). 10 proteins were found under an FDR-corrected threshold (p-value < 6.0 × 10^^-4^). Nine out of these 10 proteins were (p-value < 1.7 × 10^^-5^, Bonferroni-corrected) associated with both random glucose and pT2D. There were 484 proteins when the 10 FDR corrected protein associated with iT2D were included

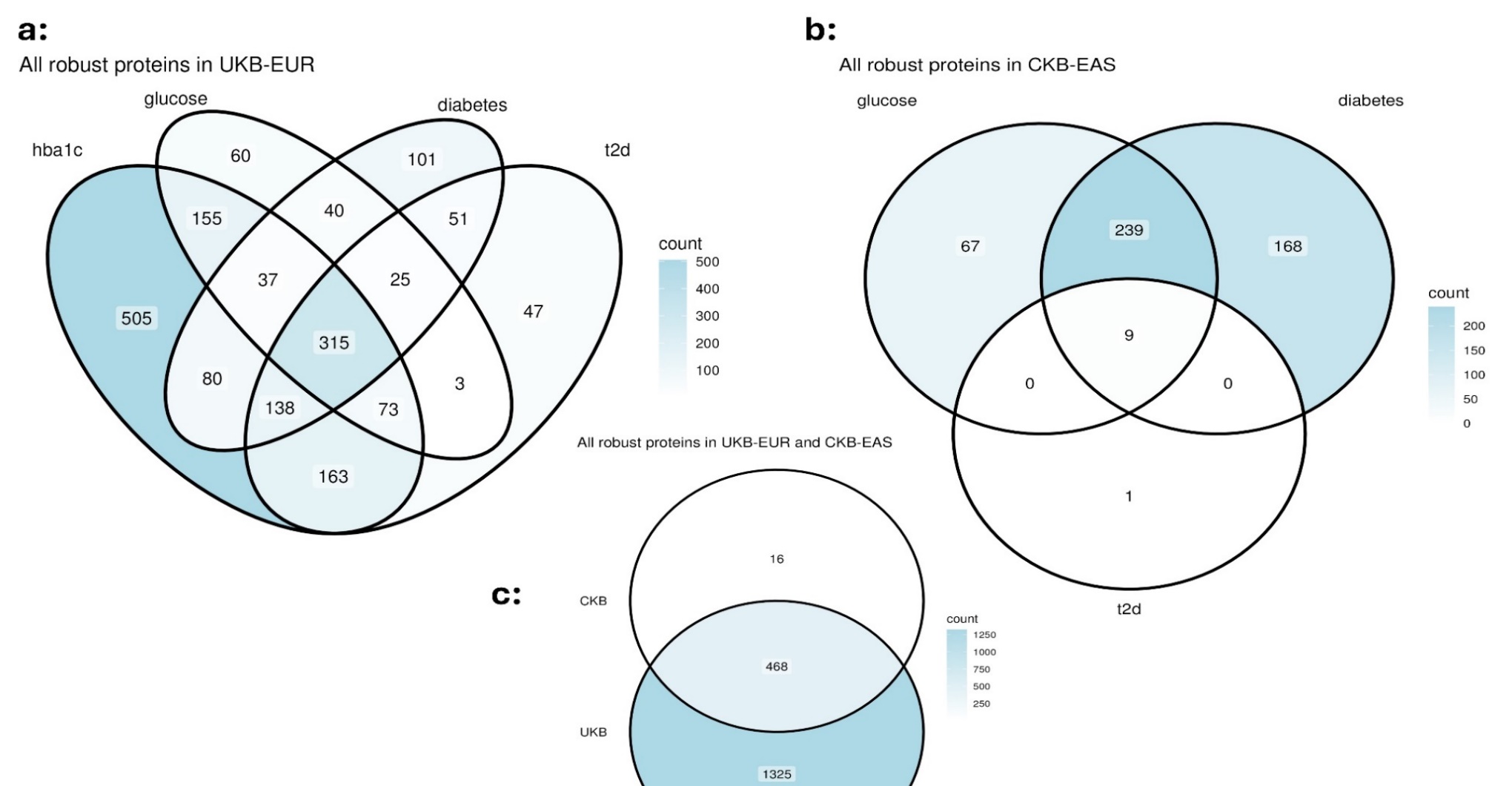

**Supplementary Fig. 3. Colocalization (stacked plot) and Mendelian randomization (forest plot) estimates of CD34, KCTD5, and SCT with T2D risk in European population**

There was no colocalization evidence of CD34, KCTD5, and SCT with T2D in EAS population; no MR analysis was conducted

CD34: Hematopoietic progenitor cell antigen CD34, KCTD5: BTB/POZ domain-containing protein KCTD5, SCT: Secretin, T2D: type 2 diabetes, IVW: inverse variance weighted, MR-PRESSO: Mendelian randomization pleiotropy residual sum and outlier; “_raw”: the MR estimates without excluding outlier genetic variants; "_outlier": the MR estimates excluding the outlier genetic variants, which were detected by the MR-PRESSO method.

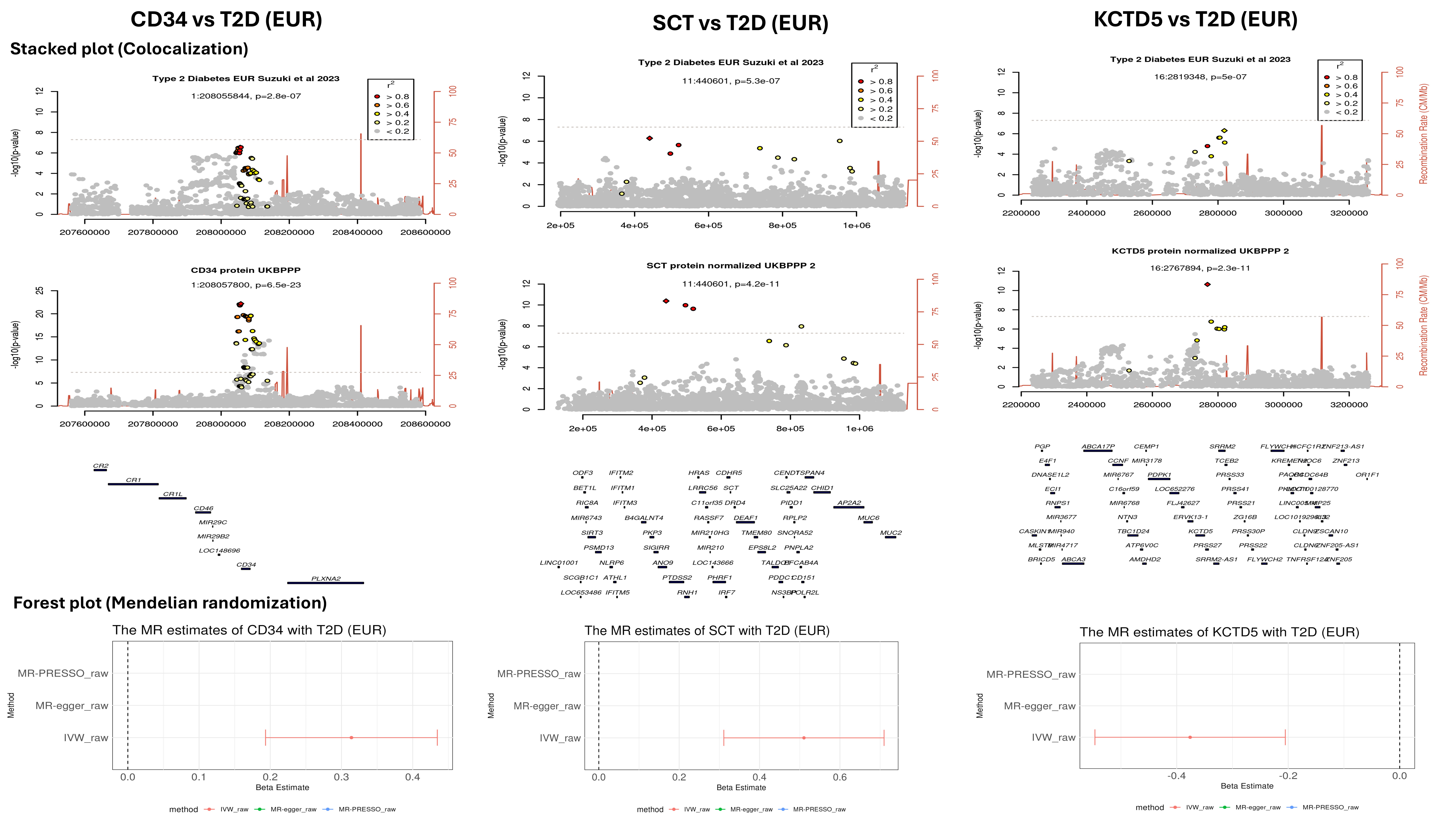

**Supplementary Fig. 4. Correlations of the observational estimates (protein–diabetes trait associations) between UK Biobank (UKB-EUR) and China Kadoorie Biobank (CKB-EAS)**

For random glucose and self-reported pT2D, the proteins highlighted in Red were those with p-value < 1.7 × 10^^-5^ (Bonferroni-corrected); for T2D incidence, the proteins highlighted in Red were those with p-value < 6.0 × 10^^-4^ (FDR corrected)

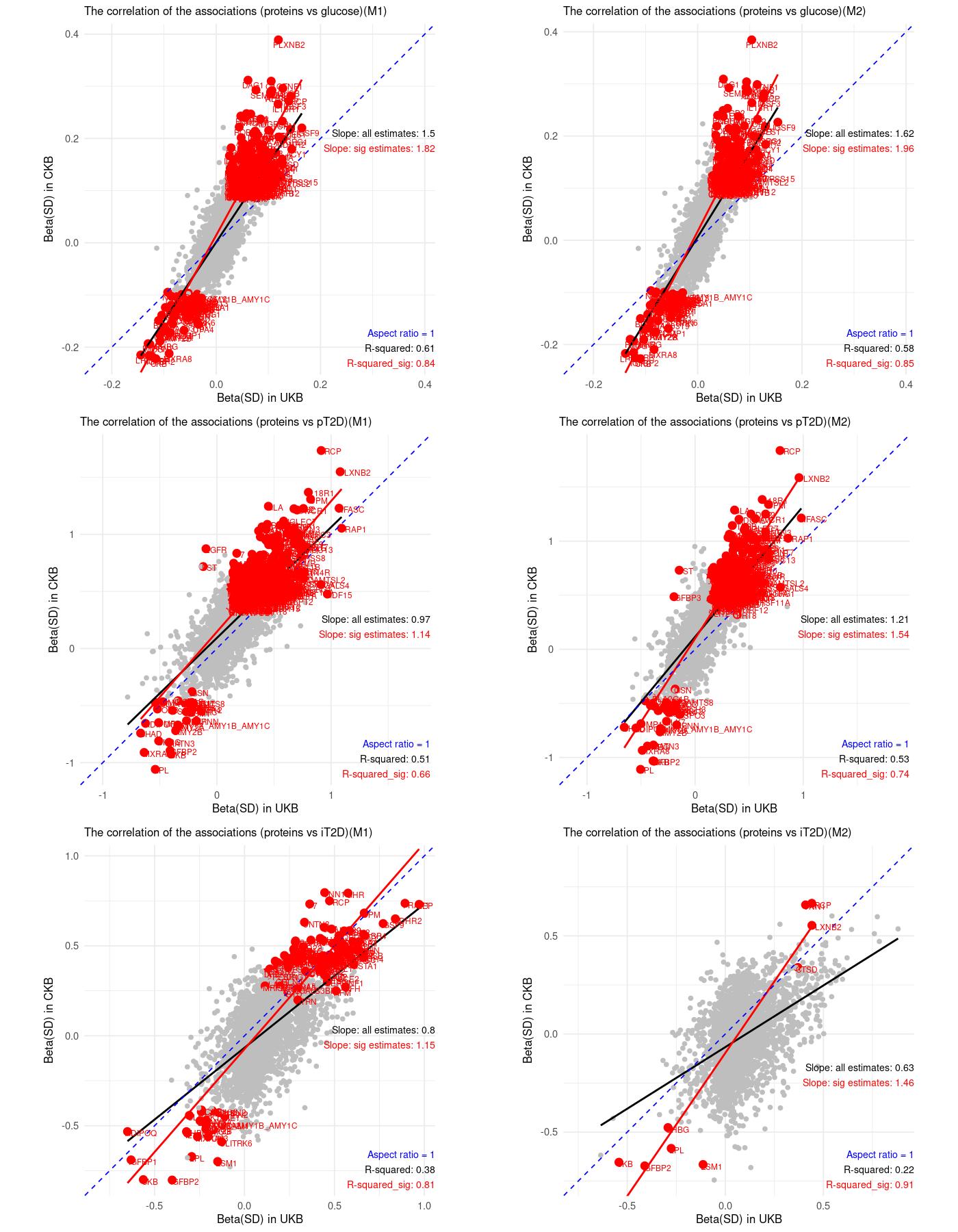

**Supplementary Fig. 5. Number of overlapped cluster-leading proteins using the leading method and the cut-off method**

Cutoff: the cutoff method: picking up the leading traits from a statistical perspective. It aggregated the weights from all the clusters and plotting them in descending order then fitted a line to the top 1% of the weights and another line to the bottom 80% of the weights. The point where the distance to the first line became shorter than to the second line was considered as cutoff for cluster weights, then the proteins were considered the most important ones.

Top10: the leading method, picking up the leading top 10% of the proteins in terms of ranking in each cluster

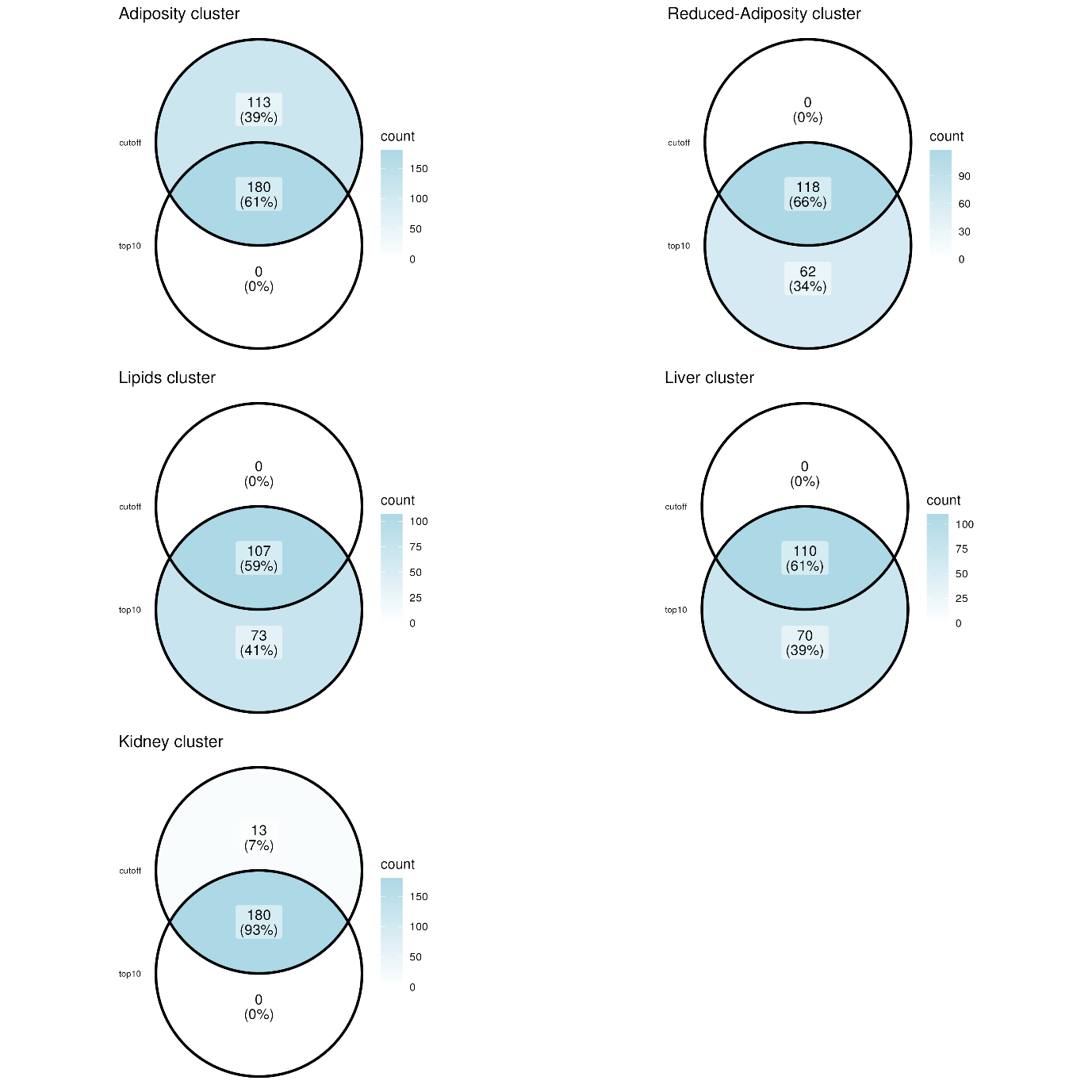
