## Supplementary Information for "Identifying molecular pathways of type 2 diabetes using proteomics, metabolic, and anthropometric profiles in UK and Chinese adults"

**UK Biobank**

### Lifestyle and environment questions

Information on lifestyle and socio-demographic characteristics were obtained using a touchscreen questionnaire at the baseline assessment. Of the lifestyle and environment questions, participants were asked about their smoking status (categorised into ‘never’, ‘former’ or ‘current’) (Data Field: 20116) and their alcohol intake frequency (categorised into ‘never’, ‘occasionally’, ‘1-3 times a month’ ‘once or twice a week’, ‘3-4 times a week’ or ‘daily’) (Data Field: 1558). The summed metabolic equivalent task (MET) score per week for all activity (Data Field: 22040) was derived according to International Physical Activity Questionnaire (IPAQ) guidelines. Participants were asked which qualifications they had. A categorial variable was generated for education (Data Field: 6138) in the UKB corresponding to 5 International Standard Classification of Education (ISCED) codes based on the years of education in UKB^1^ (5: College or university degree / NVQ or HND or HNC or equivalent; 4: Other prof.equal. eg: nursing, teaching; 3: A levels / AS levels or equivalent; 2: O levels / GCSEs or equivalent / CSEs or equivalent; 1: None of the above). Townsend deprivation index^2^ (Data Field: 22189) was calculated based on the preceding national census output areas, where each participant was assigned a continuous score corresponding to the output area in which their postcode was located. A higher index indicates a greater level of deprivation. Family history of diabetes (father, mother, sibling illness) was obtained from Data Field: 20107, Data Field:20110, and Data Field:20111. We generated three categories (at least 1 member has diabetes; at least 1 member has no diabetes but other diseases; Do not know/Not to answer/None). Other diseases included Heart disease, Stroke, High blood pressure, Chronic bronchitis/emphysema, Alzheimer's disease/dementia, Parkinson's disease, Severe Depression, Lung cancer, Bowel cancer, Breast cancer.

### Anthropometric and metabolic traits

We considered 43 baseline anthropometric and metabolic traits including BMI (Data Field: 23104), waist circumference (WC) (Data Field: 48), hip circumference (HC) (Data Field: 49), diastolic blood pressure (DBP) (Data Field: 4079), systolic blood pressure (SBP) (Data Field: 4080), whole body fat mass (WBFM) (Data Field: 23100), whole body fat-free mass (WBFFM) (Data Field: 23101), trunk fat mass (TFM) (Data Field: 23128), trunk fat-free mass (TFFM) (Data Field: 23129), mean grip strength (left (Data Field: 46) and right (Data Field: 47)), basal metabolic rate (BMR) (Data Field: 23105), forced expiratory volume in 1-second (FEV1) (Data Field: 3063), forced vital capacity (FVC) (Data Field: 3062), peak expiratory flow (PEF) (Data Field: 3064), direct low-density lipoproteins cholesterol (LDL) (Data Field: 30780), high-density lipoproteins cholesterol (HDL) ((Data Field: 30760), total cholesterol (TC) (Data Field: 30690), total triglycerides (TG) (Data Field: 30870), apolipoprotein A (Apo A) (Data Field: 30630), apolipoprotein B (Apo B) (Data Field: 30640), Lipoprotein A (Lipo A) (Data Field: 30790), sex hormone binding globulin (SHBG) (Data Field: 30830), testosterone (Data Field: 30850), oestradiol (Data Field: 30800), insulin-link growth factor 1 (IGF-1) (Data Field: 30770), alanine aminotransferase (ALT) (Data Field: 30620), alkaline phosphatase (ALP) (Data Field: 30610), aspartate aminotransferase (AST) (Data Field: 30650), gamma glutamyltransferase (GGT) (Data Field: 30730), albumin (Data Field: 30600), total bilirubin (Data Field: 30840), direct bilirubin (Data Field: 30660), total protein (Data Field: 30860), urate (Data Field: 30880), urea (Data Field: 30670), creatinine (Data Field: 30700), cystatin C (Data Field: 30720), phosphate (Data Field: 30810), rheumatoid factor (Data Field: 30820), C-reactive protein (CRP) (Data Field: 30710), Vitamin D (Data Field: 30890), calcium (Data Field: 30680)) as diabetes risk factors. Detailed information of the above traits would be referred to the UKB showcase <https://biobank.ndph.ox.ac.uk/showcase/>.

### Glycaemic traits

HbA1c was measured in red blood cells by HPLC on a Bio-Rad VARIANT II Turbo analyzer and glucose was assayed in serum by hexokinase analysis on a Beckman Coulter AU5800.^3^ Samples were assumed to be non-fasting, because participants were not advised to fast before attending. They were asked to record the last time they ate or drank anything other than water before attending the clinic and those answers were used to derive ‘fasting time’ prior to sampling. During routine quality control checks, the UKB laboratory team observed that, due to a sample processing error, ~ 8% of the glucose assay results were lower than expected and therefore a ‘dilution correction factor’ was provided and applied to these results.^4^ The HbA1c samples were not affected.

### T2D prevalence and incidence

Baseline T2D (prevalence) was considered of those who were 1) baseline reported doctor-diagnosed diabetes (Data Field: 2443): 2) whose date of first reported diabetes (ICD-10, E11: non-insulin-dependent diabetes mellitus, Data Field: 130708) ahead of date of attending assessment; 3) random blood glucose >= 11.1 mmol/l OR HbA1c >= 48 mmol/mol. It is possible to misclassify a few type 1 diabetes prevalence cases into pT2D. However, no self-reported diabetes cases were approximately diagnosed before 25 years old in the UKBPPP subsample.

T2D incidence cases were identified in according the ICD-10 code (E11: non-insulin-dependent diabetes mellitus) (Data Field: 130708).^5^ The censoring / leaving time was considered to be earliest among the following dates: 1) T2D diagnosed date; 2) censoring date from the hospital inpatient data (Data Field 40022); 3) death date (Data Field: 40000); 4) Lost to follow-up date (Data Field: 191). The censoring dates of the participants from Hospital Episode Statistics for England (HES), Scottish Morbidity Record (SMR), Patient Episode Database for Wales (PEDW) are different (HES: 31 Oct 2022, SMR 31 Aug 2022, PEDW: 31 May 2022). Besides, the censoring date of the hospital inpatient data was based on the last day of the month for which the number of records was greater than 90% of the mean of the number of records for the previous three months. In this case, some of the incidence cases occurred after the censoring date. The participants were not fixed into the same assessment centre throughout the follow-up and the source of the inpatient record was inclusive (e.g., some participants have data coded as HES|PEDW, HES|PEDW|SWR, HES|SMR, PEDW|SMR) (Data Field 40022). Therefore, we considered the censoring date determined by the source of the inpatient record rather than the baseline assessment centre, unless 1) the source of the inpatient record is missing, the baseline assessment centre is applied; 2) the inpatient record is ambiguous, we followed the priority (HES, SMR, PEDW), considering the majority of the data is from HES then SMR then PEDW); 3) if the T2D diagnosed was after the censoring date, it would be considered as a case that identified on the censoring date.

**The China Kadoorie Biobank**

The baseline survey of China Kadoorie Biobank (CKB) was established during 2004 -2008 in ten geographically diverse regions in China (five were urban regions and five were rural regions). 512,715 participants aged 30–79 years were recruited. Sociodemographic characteristics (e.g., education (highest_education), income (household_income)), lifestyle (e.g., smoking (smoking_category), alcohol consumption (alcohol_category), physical activity (met)), and personal and family medical history (e.g., family history of diabetes (father_diabetes, mother_diabetes, siblings_diabetes), self-reported health status (self_rated_health)), as well as physical and blood measurements (e.g., BMI (bmi_calc), and random blood glucose (random_glucose_x10, HbA1c was not available) were measured in the baseline survey followed by three subsequent resurveys in a ∼5% subset (in 2008, 2014, and 2021, respectively). Prior to starting the project, central ethics approvals were obtained from Oxford University and the China National Center for Disease Control and Prevention (CDC). In addition, approvals were also obtained from institutional research boards at the local CDCs in the ten regions. All participants provided written informed consent.^6,7^

### T2D prevalence and incidence

The diabetes prevalence was obtained via self-reported diabetes data (i.e., participant has history of diabetes (reported OR random blood glucose >= 11.1 mmol/L OR fasting blood glucose >= 7.0 mmol/L)) in the baseline. As above, we cannot rule out misclassification of type 1 diabetes prevalence into as pT2D, though no self-reported diabetes cases were approximately diagnosed before 25 years old in the CKB Olink subsample. We identified T2D incidence cases (ep_CKB0048_combined_ep) based on the ICD10 (E11) code (follow-up till 31 December 2018). The following information would be considered as censored/leaving whenever the date of death, the date of loss to follow-up, and the date of T2D diagnosed by 31 December 2018. The detailed information of the above-mentioned characteristics are available in the CKB data showcase: <https://www.ckbiobank.org/datashowcase/18.01/showcase_contents.html>

**Genetic variants associated with diabetes traits and metabolic/anthropometric traits**

Eight diabetes related traits (T2D,^8^ HbA1c,^9^ fasting glucose,^9^ fasting insulin,^9^ random glucose,^10^ proinsulin,^11^ modified Stumvoll insulin sensitivity index (ISI)^12^ and the fold-change in insulin concentration (IFC)^12^) genome-wide association studies (GWAS) were available for the EUR population. The summary statistics of T2D was obtained from the Type 2 Diabetes Global Genetics Initative (T2DGGI), which is, so far, the largest T2D GWAS containing 428,452 T2D cases and 2,107,149 controls (effective sample size = 1,246,658).^8^ The seven glycaemic and insulin traits were downloaded from the MAGIC (the Meta-Analyses of Glucose and Insulin-related traits Consortium). The summary statistics of HbA1c, fasting glucose (BMI adjusted, BMI unadjusted is not available), and fasting insulin (BMI adjusted, BMI unadjusted is not available) were obtained from an aggregated GWAS^9^ in up to 281,416 individuals without diabetes (N for HbA1c = 146,806, 1SD = 41%; N for fasting glucose = 200,622, 1SD = 0.83 mmol/l; N for fasting insulin = 159,304, 1SD = 0.43 natural log transformed of pmmol/l). Random glucose was also considered, where the EUR specific summary statistics were downloaded from a GWAS study with in 476,326 individuals without diabetes of European (N = 459,772) and other ancestries (N = 16,554), along with the exclusion of extreme hyperglycemia (random glucose > 20 mmol l/1) and individuals with diabetes (1 SD = 0.92 mmol/l).^10^ Summary statistics of proinsulin,^11^ ISI,^12^ and IFC^12^ were from two relative smaller sample GWASs. For proinsulin, the summary statistics (SD unit inverse normal transformation) were from 16 European-ancestry studies in 45,861 individuals, who were free from diabetes (i.e., diabetes diagnoses, a diabetes treatment, had fasting glucose ≥ 7 mmol/L, 2-h glucose >= 11.1 mmol/L, or HbA1c >= 6.5% (48 mmol/mol)). ISI and IFC were two oral glucose tolerance test (OGTT) derived measures of insulin resistance after a glucose challenge (subsequently referred to as post-challenge insulin resistance). ISI incorporates measures of insulin and glucose during an OGTT, which was to approximate hyperinsulinemic-euglycaemic measures of whole-body insulin sensitivity. Lower values of ISI reflect greater insulin resistance. IFC reveal the relationship between fasting and post challenge plasma insulin. Higher IFC values indicates more insulin is required to lower blood glucose after a standard glucose load, which indicates post-challenge insulin resistance. Summary statistics were from a GWAS study of 53,710 (ISI, SD unit inverse normal transformation) and 53,334 (IFC, SD unit inverse normal transformation) participants without diabetes from 3 ancestry groups across 28 studies.^12^

For ESA population, four diabetes related traits (T2D,^8^ HbA1c,^13^ fasting glucose,^13^ fasting insulin^9^) were available. The summary of T2D was obtained from the T2DGGI, which included 24,7158 effective sample (88,109 cases vs 339,395 controls).^8^ For the two glycaemic traits, the estimates were from a GWAS, to our knowledge the largest GWAS in EAS population, combing the Taiwan Biobank and Biobank Japan, where 163,836 for HbA1c and 225,951 for fasting glucose were assessed (SD unit inverse normal transformation).^13^ One insulin related trait was available in EAS population, fasting insulin (BMI adjusted, BMI unadjusted is not available) was assessed with the data downloaded from MAGIC, though the GWAS sample size was relative small (N = 29,792, SD unit inverse normal transformation).^9^

For EUR population, the GWAS summary statistics of 28 cluster leading metabolic/anthropometric traits were available. The summary statistics of BMI and WHR, which were obtained from a meta-analysis combining the results from the Genetic Investigation of ANthropometric Traits (GIANT) consortium and UKB (BMI: N total = 806,834, N UKB = 484,680, N GIANT = 322,154; WHR: N total = 697,734, N UKB = 485,486, N GIANT = 212,248; SD unit inverse normal transformation).^14^ The summary of others metabolic/anthropometric traits (WC, HC, WBFM. WBFFM, TFM. TFFM, BMR, pulse rate, TG, TC, LDL, HDL, Apo A, Apo B, ALT, ALP, AST, GGT, cystatin C, creatinine, urea, urate, SHBG, calcium, albumin, and total protein) were downloaded from Pan-UK Biobank (<https://pan.ukbb.broadinstitute.org/>), which releases the results of a large genome-wide analysis for a wide range of outcomes in a diverse group (e.g., different ancestries) of individuals from the UK (N EUR = 420,531; SD unit inverse normal transformation)

For EAS population, the GWAS summary statistics of 17 cluster leading metabolic/anthropometric traits (BMI, WHR, WC, HC, TG, LDL, HDL, TC, ALT, AST, ALP, GGT, creatinine, urea, calcium, albumin, total protein) as those tested in EUR were available. Two additional metabolic/anthropometric traits (body fat rate (BFR) and resting heart rate (RHR)) were applied, which were measuring the similar characteristics as WBFM and pulse rate did. The summary statistics of BMI (N = 256,450), TG (n = 204,282), LDL (N = 165,481), HDL (N = 167,585), TC (N = 228,423), ALT (N = 243,160), AST (n = 242,683), GGT (N = 226,086), creatinine (N = 242,881), urea (N = 241,382), and albumin (N = 213,154) were from the GWAS combing the Taiwan Biobank and Biobank Japan.^13^ For WHR (N = 92,615), WC (N = 92,615), HC (N = 92,615), BFR (N = 92,615), and RHR (N = 92,615), the summary statistics were only available in the Taiwan Biobank cohort.^13^ Lastly, the summary statistics of ALP (N = 118,886), calcium (N = 83,980), and total proteins (N = 133,321) were from the Biobank Japan (<https://pheweb.jp/>)^15^. All the summary statistics were in SD unit with inverse normal transformation.

Detailed description of specific studies’ design and characteristics of studies’ participants were provided in the original publications and **Supplementary Table 1**.

**Statistical methods**

### bNMF clustering

In this study, we picked up the top 10% leading proteins (call it leading method) of each cluster as the candidate proteins. In Kim et al^16^ ‘s study, they picked up the leading traits from a statistical perspective (call it cutoff method here). They aggregated the weights from all the clusters and plotting them in descending order then fitted a line to the top 1% of the weights and another line to the bottom 80% of the weights. The point where the distance to the first line became shorter than to the second line was considered as cutoff for cluster weights, then the traits / proteins were considered the most important ones. We compared the two methods with the use of 1,793 proteins *vs* 42 traits matrix model (the main model with 10 times iterations). The two methods can identify similar leading proteins (**Supplementary Fig 5**) and traits (**Supplementary Table 18**).

### Colocalization

We checked whether the diabetes associated proteins share a causal variant with the diabetes traits as well as the cluster leading metabolic/anthropometric traits in a genomic region using the hypothesis prioritisation for multi-trait colocalization (HyPrColoc), which is a Bayesian method for identifying shared genetic associations between complex traits in a particular gene region using summary GWAS results. Statistical detail was described elsewhere.^17^

We conducted two-steps colocalization to pick up the candidate proteins genetically align with diabetes risk. In the 1^st^ step of colocalization, 906 proteins obtained from bNMF were processed colocalization analysis via HyPrColoc assessing the evidence of genetic alignment with 8 diabetes traits (HbA1c, fasting glucose, random glucose, fasting insulin, proinsulin, ISI, IFC, T2D). Among the 906 proteins, five proteins are associated with two different gene regions (B4GAT1 (beta-1,4-glucuronyltransferase 1) is associated with *B3GNT1* and *B3GNT6*; CKMT1A_CKMT1B (creatine kinase U-type, mitochondrial) is associated with *CKMT1A* and *CKMT1B*; DEFB4A_DEFB4B (beta-defensin 4A) is associated with *DEFB4A* and *DEFB4B*; EBI3_IL27 (interleukin-27) is associated with *EBI3* and *IL27*; IL12A_IL12B (interleukin-12) is associated with *IL12A* and *IL12B*). As such, colocalization for these five proteins would be conducted within the two gene regions respectively. Among the 906 proteins tested, 21 proteins (ACE2, AIFM1, AMOT, CD99L2, CFP, CHRDL1, EDA2R, F9, GLA, GPR64, HS6ST2, IGBP1, IL13RA1, L1CAM, LAMP2, PLXNB3, SYAP1, SYTL4, TIMP1, VSIG4, XG) are coded in the X chromosome, which were not processed to the genetic analyses in below due to no summary statistics of the X chromosome were available in the GWAS.

In the CKB-EAS, following the same protocol as above, 878 out of 906 proteins were available. Twenty-one proteins were coded on the X chromosome (as above) and seven proteins (PALM2, AMY1A_AMY1B_AMY1C, SARG, CKMT1A_CKMT1B, DEFB4A_DEFB4B, EBI3_IL27, IL12A_IL12B) did not have pQTL in the CKB-EAS study were excluded. As such, 878 proteins were processed colocalization analysis with 4 diabetes traits (HbA1c, fasting glucose, fasting insulin, T2D). In addition, there is no available colocalization estimate of protein CD160, GDF2, and PDZK1 with T2D due to only 2 – 3 overlapped SNPs in the gene region with the T2D GWAS.

### Mendelian randomization

In UKB-EUR, the mean F-statistics of all the cis-pQTLs (protein quantitative trait loci) of the 53 (HHEX had no cis-pQLT reaching genome-wide significance) colocalized proteins were from 44 (SCT) to 393 (DNAJB6) and the R^2^ were from 0.13% (SCT) to 31.2% (SPINK4) (**Supplementary Table 7**). In CKB-EAS, the F-statistics of all the cis-pQTLs of the 9 (FST has no cis-pQLT reaching genome-wide significance) colocalized proteins were from 57 (RTBDN) to 786 (TSPAN8) and the R^2^ were from 1.4% (RTBDN) to 16.5% (TSPAN8) (**Supplementary Table 8**).

In the CKB-EAS proteomic GWAS, all the proteins tested were with one cis-pQTL and Wald ratio (WR) was applied. Whenever the cis-pQTL was not available in the outcome GWAS, a proxy with R^2^ > 0.8 would be applied. The identified proxy was only used for whenever the original cis-pQTL cannot be identified in a specific outcome. The MR estimates of the protein – trait pair would keep with the use of the original cis-pQLT whenever possible. Among the nine colocalization robust proteins (FST has no available *cis*-pQLT), proxy SNPs for two proteins (FGFBP3 and TSPAN8) were identified which solved issue due to missingness (**Supplementary Table 8**). Although proxy of ACY1 were identified, the missingness remained (**Supplementary Table 9**).

We explored screening candidate proteins via colocalization before MR, though there is no consensus of the sequency, and the two methods were not independent.^18^ We valued conducting colocalization in advance: 1) Colocalization robust could not distinguish causal effects until valid MR designs confirm the causality. Colocalization was considered a necessary condition of a causal relationship, whereas a significant MR estimate was not necessary indicate an effect, which could be due to pleiotropy and/or linkage disequilibrium (LD). 2) Colocalization leverages more genetic information regarding the different effect size, direction, genome-wide significance, and LD. 3) Colocalization was more conservative, which reduced the risk of false positive in the cis-MR design. 4) It was debatable to set up a screening criterion for MR (e.g., p-value of the Wald estimate) before processing colocalization whenever the pleiotropy robust methods were available.

### Bidirectional Mendelian randomization

Extract the instrumental variables of the diabetes traits and metabolic/anthropometric traits

We had two strategies to extract the instrumental variables (i.e., the independent SNPs in genome-wide significance) of the diabetes traits and metabolic/anthropometric traits. Whenever the independently significant SNPs were available in the discovery GWAS (i.e., specific SNPs were present discovery GWAS manuscript), we considered these SNPs were the most convincing independent instrumental variables. We called this GWAS obtained instrumental variables (“_gwas”). Otherwise, we extracted the instrumental variables following the steps below. In the full GWAS summary data, the genome-wide significance (p < 5 x 10^-8^) would be selected, and all the insertion/deletion SNPs were excluded. We then performed LD clumping (<https://mrcieu.github.io/ieugwasr/articles/local_ld.html>) locally using the *ld_clum()* function, where we set the clumping threshold r^2^ = 0.001 and using the LD reference panel of EUR from 1000 Genome reference data (EAS was applied for EAS population estimates). We called this clumping obtained instrumental variables (“_clump”). For the EUR population, the SNPs of HbA1c, fasting glucose, random glucose, fasting insulin, proinsulin, and T2D were obtained from two strategies. SNPs of other traits were from the clumping strategy. For EAS population, SNPs of T2D were from two strategies. SNPs of other traits were from the clumping strategy. As such, traits had two sets of genetic variants would have two sets of MR estimates.

### Mendelian randomization estimates Tier system

We developed a 4 levels Tier system (MRT1 – MRT4) incorporating all the MR estimates including the main (IVW) and sensitivity (MR-Egger, WM, MR-PRESSO) analyses results (**Supplementary Table 11**).

When <= 2 genetic variants are available (only the IVW estimate is available).

- MRT1 pairs are those with Bonferroni-corrected p-IVW < 0.05;
- MRT2 pairs are those with p-IVW < 0.05;
- others are classified into MRT4; MRT3 are not available in this case.

When 3 genetic variables are available, the IVW and MR-Egger estimates are available.

- MRT1 pairs are those with
  - 2-point estimates are in the same direction &
  - Bonferroni-corrected p-IVW < 0.05 &
  - Bonferroni-corrected p-Egger < 0.05
- MRT2 pairs are those with
  - 2-point estimates are in the same direction &
    - (Bonferroni-corrected p-IVW < 0.05 & p-Egger intercept >= 0.05) OR
    - (p-IVW < 0.05 & p-Egger < 0.05)
- MRT3 pairs are those with
  - 2-point estimates are in the same direction &
  - p-IVW < 0.05 &
  - p-Egger intercept >= 0.05
- MRT4 pairs are the others.

When > 3 genetic variables are available, the IVW, MR-Egger, and MR-PRESSO are available.

- MRT1 pairs are those with
  - 3 point-estimates are in the same direction &
  - (Bonferroni-corrected p-IVW < 0.05 OR Bonferroni-corrected p-PRESSO < 0.05) &
  - Bonferroni-corrected p-Egger < 0.05
- MRT2 pairs are those with
  - 3 point-estimates are in the same direction &
    - ((Bonferroni-corrected p-IVW < 0.05 OR Bonferroni-corrected p-PRESSO < 0.05) & p-Egger intercept >= 0.05) OR
    - ((p-IVW < 0.05 OR p-PRESSO < 0.05) & p-Egger < 0.05)
- MRT3 pairs are those with
  - 3 point-estimates are in the same direction &
  - p-Egger intercept >= 0.05 &
  - (p-IVW < 0.05 OR p-PRESSO < 0.05)
- MRT4 estimates are the others.

Whenever the protein – trait pair estimates were with evidence of outliers identified by MR-PRESSO, the corresponding tier was based on the outlier-corrected estimates.

In the reverse MR

When <= 2 genetic variants are available, only IVW estimate is conducted

- MRT1 pairs are those with Bonferroni-corrected p-IVW < 0.05;
- MRT2 pairs are those with p-IVW < 0.05
- others are classified into MRT4; MRT3 are not available in this case.

When >= 3 genetic variants are available, IVW, WM, and MR-Egger estimates are conducted

- MRT1 pairs are those with
  - 3 point-estimates are in the same direction &
  - Bonferroni-corrected p-IVW < 0.05 OR Bonferroni-corrected p-WM < 0.05 &
  - Bonferroni-corrected p-Egger < 0.05
- MRT2 pairs are those with
  - 3 point-estimates are in the same direction &
    - ((Bonferroni-corrected p-IVW < 0.05 OR Bonferroni-corrected p-WM < 0.05) & p-Egger intercept >= 0.05) OR
    - ((p-IVW < 0.05 OR p-WM < 0.05) & p-Egger < 0.05)
- MRT3 pairs are those with
  - 3 point-estimates are in the same direction &
  - p-Egger intercept >= 0.05 &
  - (p-IVW < 0.05 OR p-WM < 0.05)
- MRT4 estimates are the others.

For Bonferroni-corrected p values, in UKB-EUR, we considered the p value < 2.6 * 10^^-5^ (i.e., 0.05 / (53 * 36)) (HHEX has no available cis-pQTL) as statistically significant when accessing the effect of protein to trait; we considered the p value < 2.6 * 10^^-5^ (i.e., 0.05 / (54 * 36)) statistically significant when assessing the effects of traits on proteins. In CKB-EAS, we considered the p value < 2.2 * 10^^-4^ (i.e., 0.05 / (9 * 23)) (FST has no available cis-pQTL) as statistically significant when accessing the effect of protein to trait; we considered the p value < 2.2 * 10^^-4^ (i.e., 0.05 / (10 * 23)) statistically significant when assessing the effects of traits on proteins.

**References**

1.	Carter, A.R.*, et al.* Mendelian randomisation for mediation analysis: current methods and challenges for implementation. *Eur J Epidemiol* **36**, 465-478 (2021).

2.	Townsend, P., Phillimore, P. & Beattie, A. *Health and deprivation: inequality and the North*, (Routledge, 1988).

3.	UK Biobank Biomarker data. Vol. 2022.

4.	Allen, N.E.*, et al.* Approaches to minimising the epidemiological impact of sources of systematic and random variation that may affect biochemistry assay data in UK Biobank. *Wellcome Open Res* **5**, 222 (2020).

5.	Biobank, U. UK Biobank First Occurrence of Health Outcomes Defined by 3-character ICD10 code. (2019).

6.	Chen, Z.*, et al.* China Kadoorie Biobank of 0.5 million people: survey methods, baseline characteristics and long-term follow-up. *Int J Epidemiol* **40**, 1652-1666 (2011).

7.	Walters, R.G.*, et al.* Genotyping and population characteristics of the China Kadoorie Biobank. *Cell Genomics* **3**, 100361 (2023).

8.	Suzuki, K.*, et al.* Genetic drivers of heterogeneity in type 2 diabetes pathophysiology. *Nature* **627**, 347-357 (2024).

9.	Chen, J.*, et al.* The trans-ancestral genomic architecture of glycemic traits. *Nat Genet* **53**, 840-860 (2021).

10.	Lagou, V.*, et al.* GWAS of random glucose in 476,326 individuals provide insights into diabetes pathophysiology, complications and treatment stratification. *Nat Genet* **55**, 1448-1461 (2023).

11.	Broadaway, K.A.*, et al.* Loci for insulin processing and secretion provide insight into type 2 diabetes risk. *Am J Hum Genet* **110**, 284-299 (2023).

12.	Williamson, A.*, et al.* Genome-wide association study and functional characterization identifies candidate genes for insulin-stimulated glucose uptake. *Nat Genet* **55**, 973-983 (2023).

13.	Chen, C.Y.*, et al.* Analysis across Taiwan Biobank, Biobank Japan, and UK Biobank identifies hundreds of novel loci for 36 quantitative traits. *Cell Genom* **3**, 100436 (2023).

14.	Pulit, S.L.*, et al.* Meta-analysis of genome-wide association studies for body fat distribution in 694 649 individuals of European ancestry. *Hum Mol Genet* **28**, 166-174 (2019).

15.	Sakaue, S.*, et al.* A cross-population atlas of genetic associations for 220 human phenotypes. *Nature Genetics* **53**, 1415-1424 (2021).

16.	Kim, H.*, et al.* High-throughput genetic clustering of type 2 diabetes loci reveals heterogeneous mechanistic pathways of metabolic disease. *Diabetologia* **66**, 495-507 (2023).

17.	Foley, C.N.*, et al.* A fast and efficient colocalization algorithm for identifying shared genetic risk factors across multiple traits. *Nat Commun* **12**, 764 (2021).

18.	Zuber, V.*, et al.* Combining evidence from Mendelian randomization and colocalization: Review and comparison of approaches. *Am J Hum Genet* **109**, 767-782 (2022).
